# Genome sequencing of 35,024 predominantly African ancestry persons addresses gaps in genomics and healthcare

**DOI:** 10.1101/2025.10.30.25338549

**Authors:** Cecile Avery, Mojgan Babanejad, James Baker, Xavier Bledsoe, Freida Blostein, Robert W Corty, Kimberlyn Ellis, Adriana M Hung, Allison Lake, John Shelley, Quanhu Sheng, Vanderbilt University Medical Center and Alliance for Genomic Discovery Investigators, Melinda Aldrich, Melissa Basford, Lisa Bastarache, Jennifer Below, Alexander G Bick, Peter Embi, QiPing Feng, Eric Gamazon, Lide Han, Jibril Hirbo, Kayla Marginean, Jonathan Mosley, Jill Pulley, Dan M. Roden, Douglas M Ruderfer, Megan Shuey, Yu Shyr, C Michael Stein, Colin Walsh, Consuelo Wilkins

## Abstract

Genetic variation is crucial in human development, disease susceptibility, and drug response. Despite populations of African descent having the highest degree of genetic variation, genetic research has predominantly focused on populations of European descent, limiting potential for discovery. Recent studies of individuals with African ancestry have driven key medical advances benefiting all populations. Large scale, electronic health record (EHR) linked biobanks have provided opportunities to expand genetic research to larger and more diverse populations. We present initial results from the Alliance for Genomic Discovery (n = 35,024), in which 80% of participants have majority African ancestry, with genome sequencing and extensive phenotyping (median ∼10 years of EHR data). We demonstrate that genetic variants known to disproportionately cause disease in patients of African descent are under-documented in the medical record, including treatable genetic conditions such as transthyretin amyloidosis. Our findings confirm that many disease-associated genetic variants have consistent effects between groups with majority African and European ancestry, including common variation, structural variation, and clonal hematopoiesis of indeterminate potential. We discover novel variants associated with drug adverse events and diagnostic codes, powered by the increased frequency of these variants in individuals with majority African ancestry. Furthermore, we report independent effects of genetic risk and social factors on glycemic control in individuals with type 2 diabetes. Overall, this work highlights the value of integrating genome sequencing and deep phenotyping in genetically diverse populations to broaden our understanding of human health and disease.

## Introduction

Populations of African descent have the highest degree of genetic variation and thus present the greatest opportunity to link genetic variation to diseases and traits^1,2^. This benefit is exemplified by the discoveries of loss-of-function variants that led to PCSK9 inhibitors^3^, the role of *G6PD* variants in diabetes complications^4^, and the association between APOL1 and chronic kidney disease^5^, all of which would not have been possible had it not been for research participants from populations of African descent^6^.

Most genetic research, however, has been conducted among individuals of European descent, limiting the potential benefits of genomic research for all populations. Large-scale EHR-linked biobanks have expanded the power of genetic studies but include primarily persons with predominant European ancestry^7–11^. Recently, research programs such as All of Us (AoU) and the Million Veterans Program (MVP) have made significant contributions to advancing genomic resources for individuals from understudied genetic ancestries^12,13^. However, even in these enriched cohorts, individuals of African ancestry represent less than 25% of the entire sample (19% in MVP and 23% in AoU).

Here, we present initial results from the Alliance for Genomic Discovery (AGD), an academic-industry partnership that generated whole genome sequence data in Vanderbilt University Medical Center’s deeply phenotyped, EHR-linked biobank (BioVU)^14^. In this work, we describe findings from 35,024 individuals (“AGD35k”), selected such that 80% of the sample is persons with majority African genetic ancestry. Our findings address multiple clinically relevant scientific questions with broad implications for the health of African and non-African populations. We provide evidence that genetic diseases are clinically under-documented including some that are treatable and that social factors contribute significantly to symptom control in diabetes. We further identify novel genetic associations that are discoverable because risk variants are present in higher frequencies in individuals with majority African ancestry. The extensive genetic and clinical information for the AGD35k’s majority African ancestry population has enabled opportunities to shed light on links between genetic variation and clinical outcomes, highlighting areas to improve intervention.

## Results

### Characterization of sample selection, demographics, and ancestry

Of the 35,024 participants from the Vanderbilt University Medical Center (VUMC) EHR-linked biobank (BioVU) selected for whole genome sequencing, the AGD35k cohort includes 34,963 participants that met inclusion criteria for this analysis. Participants had a median age of 45 years at last documented International Classification of Diseases (ICD) Clinical Modification billing code, and 61% had an EHR-reported sex of female (**Table 1**). The AGD35k cohort was selected using a propensity score approach designed to enrich the sample for participants with relative majority African ancestry (**see Methods**). Briefly, the propensity scores were calculated from a subsample of participants with known majority African ancestry based on existing genome-wide array data using EHR-reported race and ethnicity as predictors. We recognize that race and ethnicity are social constructs that correlate with but do not represent genetic ancestry. Sample selection for the AGD35k cohort was intended to enrich for those of African ancestry knowing this was an imperfect model. Most of the AGD35k cohort (n = 27,782) had majority (>50%) African ancestry when projected into the principal component space of the merged 1000 Genomes Project and Human Genome Diversity Project which includes five categories of overall continental ancestry for proportional associations (**Figure 1A-C**, **Table 1**). Among the samples selected without prior knowledge of their African genetic ancestry (n=21,653), 30% had majority non-African ancestry of which 90% had “Unknown” or “Other” EHR-reported race.

**Figure 1:**
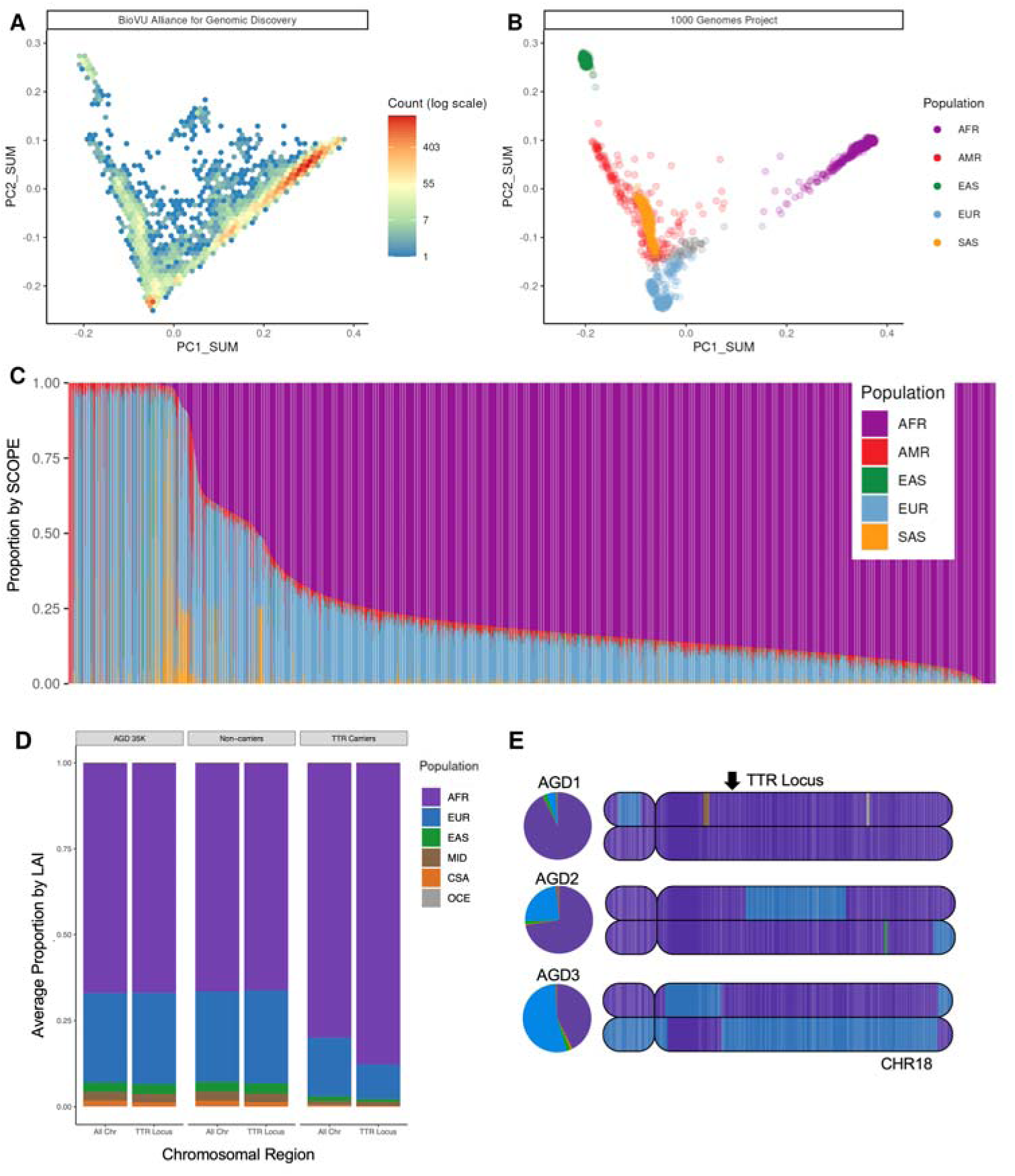
Global and local ancestry inference (LAI) in the AGD35k cohort. **A)** Density plot of the BioVU Alliance for Genomic Discovery (AGD35K) participants genomic data projected into 1000 Genomes Project (1kGP + Human Genome Diversity Project (HGDP) principal component space. **B)** 1kGP participant data projected into the same principal component space (AFR – African, AMR – American, EAS – East Asian, EUR – European, SAS – South Asian). **C)** Ancestry proportion estimates of BioVU AGD35k participants calculated using supervised SCOPE analysis and allelic frequencies from the 1kGP. Each bar represents a single participant, and bars are ordered by the proportion of African ancestry. **D)** Proportion of participants genome by continental ancestry based on LAI from the 1kGP + HGDP reference panel (AFR- African, EUR- European, EAS- East Asian, MID- Middle Eastern, CSA- Central South Asian, OCE- Oceanic) for TTR pathogenic variant carriers, non-carriers and the total AGD35k population. **E)** Painted karyograms for chromosome 18 from three representative TTR carriers (V142I). Arrow annotates the TTR locus. LAI-derived global ancestry proportions for each participant are represented to the left of the karyograms as pie charts. AGD1 and AGD2 represent two TTR carriers with varying, but majority AFR global ancestry (92.6% and 72.5%, respectively). AGD3 represents a TTR carrier with majority EUR global ancestry (53.8%) and AFR ancestry at the TTR locus.

**Table 1:** Sociodemographic, medical, and genetically inferred ancestry variables among 34,963 BioVU participants in the Alliance for Genomic Discovery sample.

| Characteristic | N = 34,963<sup>1</sup> |
| --- | --- |
| Age at last ICD code<sup>2</sup> | 45 (28, 61) |
| EHR-reported sex |  |
| Female | 21,187 (61%) |
| PLINK sex |  |
| Female | 21,182 (61%) |
| Excluded | 15 |
| EHR-reported race |  |
| American Indian/Alaskan Native | 31 (<0.1%) |
| Asian | 120 (0.3%) |
| Black | 28,523 (82%) |
| Hispanic | 248 (0.7%) |
| Unknown | 2,586 (7.4%) |
| White | 1,263 (3.6%) |
| All others (multiple race categories, other, etc) | 2,192 (6.3%) |
| EHR record length, years<sup>2,3</sup> | 10 (4, 17) |
| Count of unique phecodes<sup>4</sup> | 49 (24, 92) |
| Count of unique ICD entry dates<sup>3</sup> | 42 (16, 96) |
| Ancestry mixture proportions |  |
| African ancestry proportion<sup>5</sup> | 0.82 (0.67, 0.88) |
| American ancestry proportion<sup>5</sup> | 0.016 (0.003, 0.030) |
| European ancestry proportion<sup>5</sup> | 0.14 (0.08, 0.25) |
| East Asian ancestry proportion<sup>5</sup> | 0.002 (0.000, 0.008) |
| South Asian ancestry proportion<sup>5</sup> | 0.00 (0.00, 0.01) |
| Majority ancestry determination<sup>5</sup> |  |
| Admixed (majority ancestry < 0.5) | 986 (2.8%) |
| African | 27,782 (79%) |
| American | 604 (1.7%) |
| East Asian | 180 (0.5%) |
| European | 5,130 (15%) |
| South Asian | 281 (0.8%) |
| Obesity (phecode 278.1)<sup>4</sup> | 10,357 (31%) |
| Tobacco use disorder (phecode 318)<sup>4</sup> | 7,586 (23%) |
| Hyperlipidemia (phecode 272.1)<sup>4</sup> | 10,055 (30%) |
| Essential hypertension (phecode 401.1)<sup>4</sup> | 15,412 (47%) |
| Amyloidosis (phecode 270.33)<sup>4</sup> | 238 (0.7%) |
| Sickle cell trait or anemia (phecode 282.5)<sup>4</sup> | 1,151 (3.5%) |
| Type 2 Diabetes<sup>6</sup> | 6,919 (20%) |
| ACS-derived financial insecurity metric | 0.12 (0.07, 0.19) |
| Unknown | 4,744 |
| NLP-derived financial insecurity | 9,619 (28%) |

<sup>1</sup> Median (Q1, Q3); n (%)
<sup>2</sup> 66 individuals without any ICD 9/10 codes
<sup>3</sup> Calculated as the time difference between the first and last ICD code
<sup>4</sup> 1,952 individuals did not meet the phecode cohort data floor inclusion criteria, i.e. 3 unique dates upon which ICD codes were entered, and 30 days between first ICD code and last ICD code. These individuals excluded from denominator when calculating prevalence/percentages. Prevalence is of at least one ICD code within phecode definition.
<sup>5</sup> Ancestry proportion estimates derived using supervised SCOPE ancestry estimations and allelic frequencies from the 1000 Genomes Project reference populations. Categorization based on continental ancestry proportion which exceeded 0.5, individuals with no continental ancestry proportion exceeding 0.5 were considered admixed.
<sup>6</sup> Type 2 Diabetes was defined as having two outpatient diabetes codes on different dates, one diabetes code, and a qualifying HbA1c > 6.5, or having a diabetes code and being on diabetes medication. Patients with gestational diabetes or only one diabetes code were excluded. The date of the first diabetes code was considered the date of diabetes diagnosis and was required to occur at age 18 or older.
ICD = International classification of Disease codes; EHR = Electronic health record; ACS = American Community Survey; NLP = Natural language processing

**Table 2:** Sociodemographic and clinical variables among a total of 5,661 individuals with majority African ancestry in the diabetes cohort with available hemoglobin A1c and/or glucose measurements.

| Characteristic<br>(Combined N=5,661) | HbA1C<br>(N=4,459) | Glucose<br>(N=5,631) |
| --- | --- | --- |
| **Phenotype criteria met** |  |  |
| Two outpatient DM codes | 3770 (84.5%) | 4352 (77.3%) |
| One DM code and on medication | 304 (6.8%) | 807 (14.3%) |
| One DM code and one HbA1c > 6.5 | 385 (8.6%) | 472 (8.4%) |
| **Genetically determined sex** |  |  |
| Female | 2567 (57.6%) | 3204 (56.9%) |
| Male | 1892 (42.4%) | 2427 (43.1%) |
| **G6PD deficiency trait** |  |  |
| No | 3658 (82.0%) | 4607 (81.8%) |
| Yes | 801 (18.0%) | 1024 (18.2%) |
| **Proportion of census tract below poverty level** |  |  |
| Mean (SD) | 0.14 (0.10) | 0.14 (0.10) |
| Median (Min, Max) | 0.12 (0.01, 0.80) | 0.12 (0.01, 0.80) |
| Missing | 586 (13.1%) | 753 (13.4%) |
| **NLP-derived financial insecurity** |  |  |
| No | 2151 (48.2%) | 3033 (53.9%) |
| Yes | 2308 (51.8%) | 2598 (46.1%) |
| **Diabetes medication<sup>1</sup>** |  |  |
| No | 1400 (31.4%) | 1897 (33.7%) |
| Yes | 3059 (68.6%) | 3734 (66.3%) |
| **Age at first post-diagnosis measurement** |  |  |
| Mean (SD) | 54.09 (13.06) | 54.22 (13.14) |
| Median (Min, Max) | 54.43 (18.40, 89.95) | 54.88 (18.40, 89.88) |
| **Body mass index (730 days prior to DM diagnosis)** |  |  |
| Mean (SD) | 34.86 (9.15) | 34.48 (9.12) |
| Median (Min, Max) | 33.57 (13.94, 88.95) | 33.27 (13.94, 88.95) |
| Missing | 921 (20.7%) | 1149 (20.4%) |
| **eGFR (730 days prior to DM diagnosis) <sup>2</sup>** |  |  |
| Mean (SD) | 72.20 (30.82) | 71.04 (31.15) |
| Median (Min, Max) | 76.29 (1.91, 149.40) | 74.99 (1.91, 149.40) |
| Missing | 1295 (29.0%) | 1675 (29.7%) |
| **Median laboratory value <sup>3</sup>** |  |  |
|  | *Unit = %* | *Unit = mg/dL* |
| Mean (SD) | 7.23 (1.76) | 133.75 (49.06) |
| Median (Min, Max) | 6.75 (3.80, 15.90) | 119.00 (44.00, 375.00) |

1 Presence of any diabetes medication during 1 year prior to first measurement
2 Estimated Glomerular Filtration Rate (GFR) using race-free CKD-EPI equation
3 Median lab value within the 1 year follow up period after the first measurement
Abbreviations: DM, Diabetes Mellitus; HbA1c, hemoglobin A1c; G6PD, glucose-6-phosphate dehydrogenase; SD, standard deviation; NLP, natural language processing

Continental ancestry metrics provide a simple consolidation of an individual’s genetic ancestry proportions, but do not capture the complex distribution of inherited ancestral regions distributed across the genome. We utilized local ancestry inference (LAI) to provide important context to the genetic variation that is present by decomposing admixed chromosomal regions and identifying their ancestral origins (**see Methods**). As expected, we found that individuals with similar levels of African genetic ancestry can have vastly divergent local ancestry patterns across the genome. Furthermore, this difference in local ancestry composition can be clinically relevant, when disease-associated variants occur on different ancestral haplotypes at varying frequencies. One example of this phenomenon is hereditary transthyretin amyloidosis (hATTR, MIM:105210), a rare genetic disease known to disproportionately affect individuals of African descent almost entirely due to a single pathogenic variant (V142I) in the *TTR* gene seen at higher frequencies in this population. Within a set of 1,108 carriers of pathogenic variants in the *TTR* gene (1,102 had V142I, described further below), there was a significant enrichment of local African ancestry haplotypes at the *TTR* gene locus, compared to non-carriers (**Figure 1D**, p < 2x10^-16^, beta=1.313, SE=0.066). Although the carriers were also enriched for global African ancestry proportions (mean: 80.10% compared to 66.53% in non-carriers, p < 2x10^-16^, beta=2.8831, SE=0.2073), the proportions were highly variable ranging from 4.81% to 99.55% (**Figure 1E**).

AGD35k participants had rich longitudinal EHR data (median of 10 years and 69% had greater than 5 years of medical records), including information regarding medical conditions, procedures, medications, and clinical laboratory tests (**Table 1**, Figure S1). Chronic conditions were common in this cohort including obesity (31%), hypertension (47%), and hyperlipidemia (30%) (Table 1).

### Undocumented expression of disease relevant genetic variation

Four genetic conditions known to disproportionately affect individuals with higher proportions of African ancestry were selected to quantify clinical documentation related to the expression of genetic variation. Pathogenic genotypes were relatively common (**Figure 2A**): APOL1-mediated kidney disease (MIM:612551; n = 2,233), G6PD deficiency (MIM:305900; n = 1,549 including 248 homozygous females), hATTR (MIM:105210; n = 1,108) and sickle cell disease (SCD (MIM:603903; n = 240). In addition to these pathogenic genotypes, we identified 2,150 participants with sickle cell trait (SCT).

**Figure 2:**
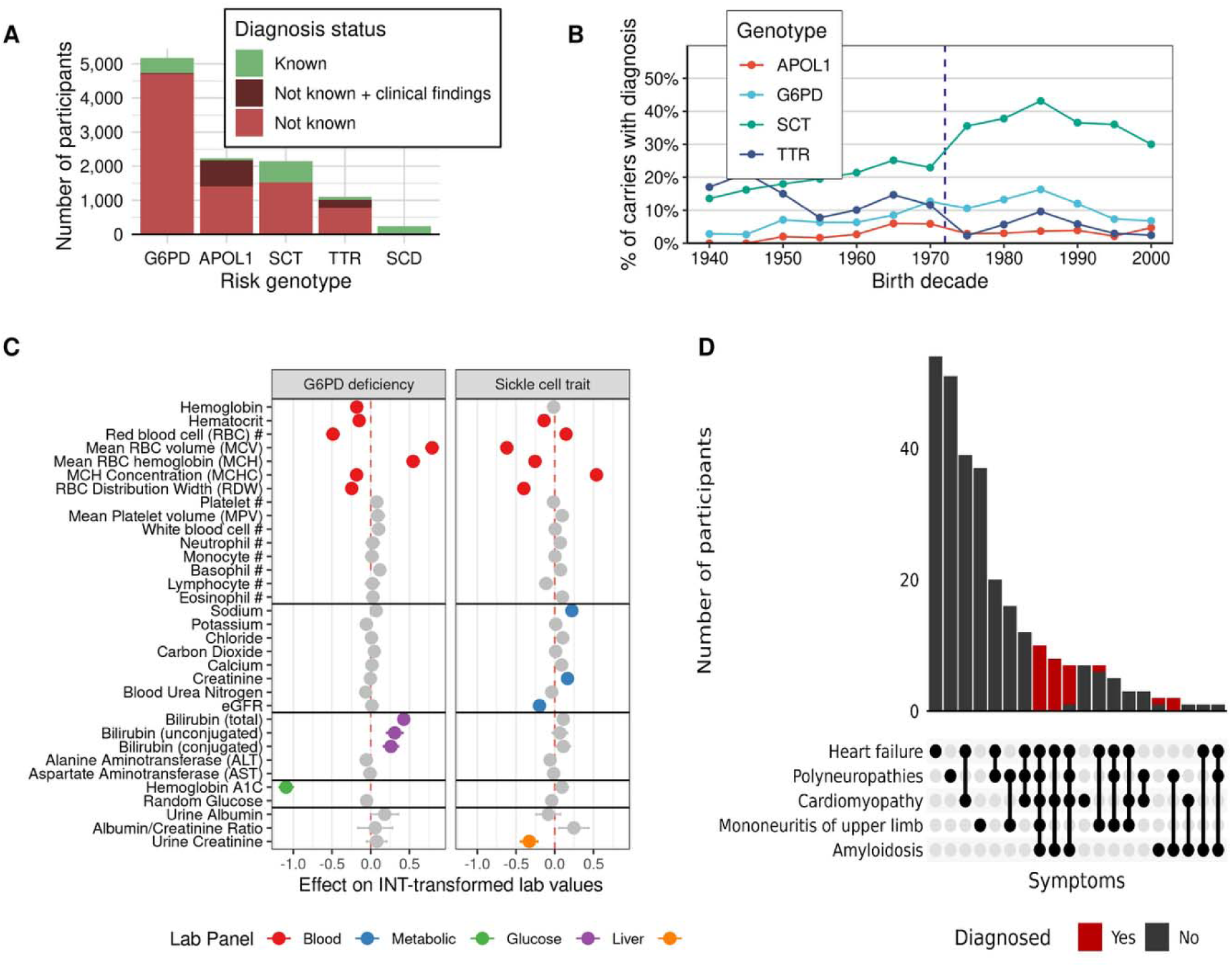
Frequency and characteristics of patients with genetic risk for APOL1, sickle cell trait, G6PD, ATTR, and sickle cell disease. **A)** Frequency of patients with known disease, patients with clinical findings relevant to the disease but lacking a diagnosis, and undocumented disease. **B)** Proportion of recognized risk carriers by participant year of birth. The vertical line depicts the passage of the National Sickle Cell Anemia Control Act of 1972 in the United States. **C)** Forest plot illustrating the effect of G6PD and sickle cell trait on commonly measured laboratory measurements among participants of majority African ancestry. Lab values were inverse-normal transformed prior to analysis. Lab effects that were significant at the Bonferroni-corrected significance threshold (*p* = 1.9×10^-4^) were colored according to the lab’s corresponding testing panel. **D)** Distribution of phenotypic heterogeneity observed in individuals with genetic risk for ATTR, colored by diagnosis status.

Diagnosis was determined by querying free text clinical documentation and billing data including diagnosis (ICD) and procedure (Current Procedural Terminology/Healthcare Common Procedure Coding System) codes (**see Methods**). EHRs for all participants who were homozygous for the sickle cell variant carried the clinical diagnosis, compared to 28.7% of participants with SCT. This rate is similar to a previous study of the UK Biobank which showed that only 22% of participants with SCT had clinical documentation^15^. EHR diagnosis rates were lower for other conditions: 9.6% for G6PD, 9.5% for hATTR, and 3.0% for APOL1. In our study, temporal trends in attribution were noted with higher EHR documentation of SCT in participants born after the National Sickle Cell Anemia Control Act of 1972, which provided federal funding for newborn screening programs (**Figure 2B**).

While under-recognized, SCT and G6PD deficiency are thought to be largely benign except for rare complications like drug-induced hemolytic anemia in G6PD deficiency. We tested whether undocumented carriers were more likely to have certain diagnoses or changes in clinically measured biomarkers compared to participants without the genotype. Among participants with majority African ancestry, SCT was not associated with any phecode after excluding participants with clinical recognition (**Table S2**). However, unrecognized SCT was significantly associated with differences in red blood cell (RBC) parameters compared to non-carriers including lower within-RBC hemoglobin content (MCH; -0.25 [-0.32 to –0.19]; P = 5.4×10^-13^) but a higher mean corpuscular hemoglobin concentration (MCHC; +0.54 [+0.47 to +0.61]; P = 2.7×10^-65^) (**Figure 2C**). SCT was also associated with a pattern of labs consistent with increased predisposition to a urinary concentration defect including lower urine specific gravity (-0.58 [-0.64 to -0.52]; P = 4.1×10^-74^), replicating previous studies^16,17^. G6PD deficiency was associated with small but significant differences in RBC parameters including higher MCH (+0.55 [+0.47 to +0.62]; P = 1.1×10^-45^). In addition, both conjugated (+0.26 [+0.17 to +0.36]; P = 6.5×10^-8^) and unconjugated bilirubin (+0.31 [+0.21 to +0.41]; P = 2.9×10^-9^) were significantly higher in G6PD deficiency carriers. This pattern of lab associations is consistent with hemolytic anemia which is the major clinical manifestation of G6PD deficiency, but the G6PD-associated hemolysis is typically acute and provoked by medication-related exposures^18^. Because we restricted to individuals with clinically unrecognized G6PD deficiency, it is possible that these asymptomatic individuals are having low-grade hemolysis that is too subtle to be detected clinically. The most important clinical implication of this low-grade hemolysis is likely the impact on estimation of glycemic control^4,19^.

Across both genotypes, the strongest lab association was the association between G6PD deficiency and hemoglobin A1c (-1.049 [-1.18 to –1.00]; P=1.4×10^-128^), with G6PD deficiency alone explaining 6.8% of the variance in A1c. There was no concomitant association with higher blood glucose. We observed that patients with unrecognized G6PD deficiency had lower odds of prediabetes (odds ratio [OR] = 0.53 [0.43 to 0.65]; P = 2.6×10^-9^) and type 2 diabetes (0.86 [0.80 to 0.92]; P = 3.2×10^-5^). Because G6PD deficiency does not impact metabolism, it is likely that these effects are entirely mediated by the inaccuracy of glycemic control estimated using A1c in these participants.

hATTR is a rare progressive disease affecting multiple organ systems for which four targeted therapies have been introduced since 2019. In individuals with *TTR* pathogenic variants over the age of 30 years old with at least 2 visits (n=802), we extracted diagnostic codes for heart failure, cardiomyopathy, amyloidosis, polyneuropathy, and carpal tunnel syndrome to capture well-established features of hATTR. In this cohort, 34% (n=272) had at least one hATTR-associated clinical feature, consistent with the incomplete penetrance associated with V142I, which consists of 98% of the *TTR* variants observed (**Table S1**)^20^. This was higher than the proportion of genotype negative controls over the age of 30 with at least one hATTR-associated feature (29%). hATTR-associated features such as amyloidosis were considerably more enriched in *TTR* carriers versus non-carriers (OR = 5.45, p<.0001) relative to more common features, such as heart failure (OR = 1.43, p<.0001).

Manual chart review of clinical notes confirmed only 29 had received a diagnosis of hATTR, with 24 having a confirmatory genetic test. For those 29 individuals, the median time from first mention of *TTR* amyloidosis to diagnosis was 49 days (IQR 10 days-296 days). 86% (n=25) of diagnosed cases had three to five of the selected diagnostic codes in their EHR (**Figure 2D**).

Clinical procedures related to the features of hATTR, such as EKGs and cardiac MRIs, were not enriched in *TTR* pathogenic variant carriers without a diagnosis compared to ancestry matched non-carriers, reflecting that these individuals do not receive additional work-up for an underlying pathology related to their genetic risk. The population attributable fraction, e.g. proportion of population with heart failure and amyloidosis due to pathogenic TTR variants in AGD35k, was 1.01% and 12.65% respectively. However, it should be noted that amyloidosis codes are not specific to hATTR and include wild type *TTR* and primary amyloidosis, for example.

### Differences in genetic effect across ancestral populations

We characterized the frequency and effect of clinically associated genetic variants in our cohort across different types of variation including somatic, structural (supplementary results), and single nucleotide polymorphisms (SNPs). From the class of somatic variants, we examined the frequency of clonal hematopoiesis of indeterminate potential (CHIP), an asymptomatic condition that is common in older adults and confers an increased risk for blood cancer, cardiovascular disease, and death^21^. CHIP was detected at a frequency that rises with age, consistent with other large studies (**Figure 3A**)^22^. CHIP was rare among people under 40 years old (< 1%) and common among people over 70 years old (>5%). Rates of CHIP were similar between individuals with majority African and European ancestry across ages (**Figure 3A**). CHIP can be caused by somatic mutations in any of ∼70 leukemia-associated genes, and the rate of CHIP driven by mutation in each gene was also similar across ancestry groups (**Figure 3B**)^21^. A genome-wide association study (GWAS) of CHIP status revealed a single, genome-wide significant association with lead SNP rs7624945 at chr3:46133575 (-log10(*p*) = 7.59, beta = 1.9) (**Figure 3C**). The minor allele G was present in ∼1-2% of persons with majority African ancestry in AGD35k and gnomAD and ∼100x rarer among all other ancestry groups in both cohorts. This variant, ∼68kb upstream of *CCR1*, has scant annotation and the association did not replicate when tested in ancestry-specific GWAS in the All of Us cohort.

**Figure 3:**
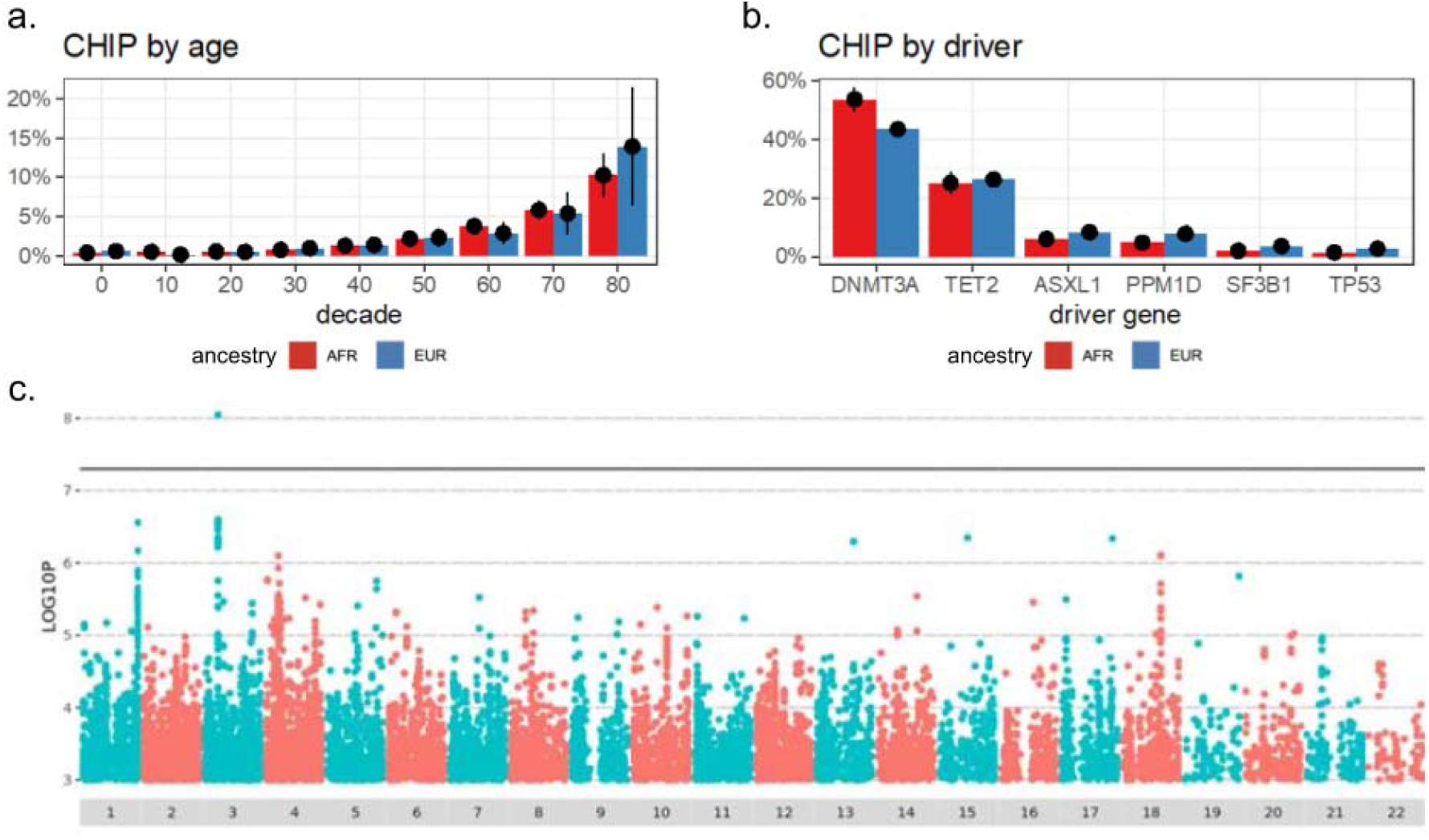
Variants of medical significance in AGD35k. **A)** The rate of CHIP by age group among people of majority African ancestry (red) compared to people of majority European ancestry (blue). Observed rate of CHIP +/- 95% confidence interval is shown. **B)** The breakdown of CHIP driver mutations in each of the five genes that most commonly harbors CHIP driver mutations among persons of majority African ancestry (red) and majority European ancestry (blue). Observed fraction of each driver gene +/- 95% confidence interval is shown. **C)** Genome-wide association study for CHIP status.

To assess the relationship between common variants and disease, we performed GWAS of 122 common diagnostic conditions selected for being seen in at least 10% of the African ancestry participants. At genome-wide significant threshold (p < 5x10^-8^), there were 63 SNP-phecode associations across 57 unique SNPs and 51 different phecodes after filtering (see Methods). Of the 63 associations, 14 (22%) were previously identified as genome-wide significant or replicated in the GWAS catalog^23^, UK Biobank^24^, FinnGen^25^ or in the European ancestry sample of AGD35k (**Table S3**). Nearly 30% of the SNPs (17 out of 57), had never been associated to any phenotype across those resources. For the 34 associations where the European ancestry minor allele frequency was greater than zero, we observed significant correlation in effect sizes between association results across African and European ancestries (r=0.42, p=0.013, Pearson). Finally, we assessed the replication rate of prior genome-wide significant association results by ancestry^26^, we replicated 6 of the 7 (85.7%) associations that were previously reported in African ancestry discovery cohorts in the GWAS catalog and for which we had sufficient power.

Comparatively, 33 of 103 (32%) associations discovered in European ancestry GWAS replicated in the African ancestry cohort of AGD35k.

### Novel genetic effects in response to drug exposure

Adverse drug events (ADEs) are among the most common preventable causes of morbidity and mortality and cost more than $100 billion annually in the US^27–29^. An estimated 20-30% of ADEs requiring hospitalization have a prominent genetic component, and the frequency of some ADEs (e.g., ACE inhibitor induced angioedema, G6PD related hemolysis) is well-known to vary by ancestry^30,31^. Further, among individuals with the same ADE, the genetic underpinnings can vary by ancestry; carbamazepine-related Stevens-Johnson syndrome is such an example^32,33^.

To discover novel genetic associations with ADEs, we restricted the AGD35k cohort to those with majority African ancestry, who had at least one problem list entry in their EHR, and who passed quality control (n = 25,130; **see Methods**). ADEs were defined as any mention of an allergy or intolerance to a medication in the problem list^34^. As part of this high throughput approach, controls for each drug/drug class were participants without mention of that drug/drug class in the problem list. The median [IQR] number of problem list entries was 3 [1, 16]. Among these patients, we identified 11 common drugs/drug classes with ADEs (**Table S4**), and validated using a sample of ≥1000 notes for each (**see Methods**). The number of drugs/drug classes for which a given participant experienced an ADE ranged from 0-7; 8,888 (35.4%) participants had a documented ADE for at least one drug/drug class (**Table S5)**. For all drugs/drug classes except aspirin, the total number of cases and controls was 25,130. For aspirin, we further excluded 2,141 participants due to potential misclassification related to a listing of aspirin because of a comorbidity contraindicating use rather than an ADE or to a listing of drug combinations (**see Methods**), resulting in a total of 22,989 cases and controls. The drugs with the most frequently documented ADEs in the problem list were penicillin (14.1%) and opioids metabolized by CYP2D6 (10.7%). All documented ADE frequencies among individuals with majority African ancestry were similar to previous reporting for individuals of African descent^35^; most of the documented ADE frequencies were lower than those previously reported among individuals with majority European ancestry, except aspirin (which was similar) and ACE inhibitors, which were higher in the African ancestry group (6.4% vs. 2.4%) (**Figure 4A**).

**Figure 4:**
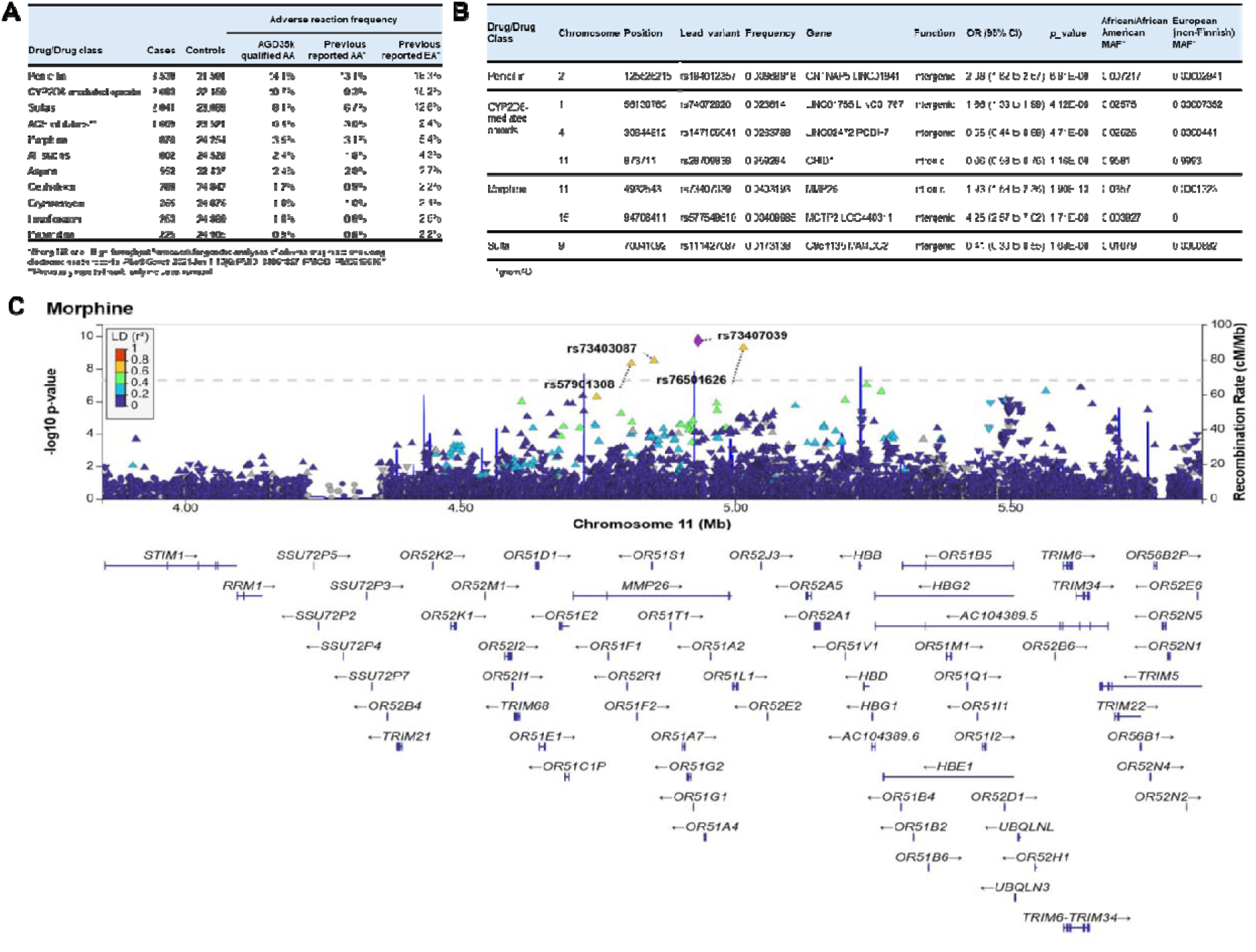
Adverse Drug Events (ADE) among the AGD35k cohort with majority African ancestry, with problem list entries, and passing quality control (n = 25,130). **A)** Frequency of Cases and Controls for ADE, compared to previously reported results. **B)** Genome-wide significant results, lead variant reported. **C)** LocusZoom Plot with *MMP26* significant results for ADE with morphine use.

A GWAS of each drug/drug class identified 7 independent genome-wide significant loci (p < 5x10^-8^) for penicillin, CYP2D6-related opioids, morphine, and sulfas (**Figure 4B**). One of the lead SNPs associated with morphine ADEs (OR=1.93 [1.58, 2.36], p=1.90 x10^-10^) is located in *MMP26* (rs73407039). This SNP has been associated with lymphocyte count^36^ (**Figure 4C**); additionally, it is in high LD (r2=0.77 in African populations) with rs112902560, which was significantly associated with levels of cystatin-C, a biomarker indicative of renal function, in a previous large GWAS^37^. In observing the LocusZoom of the *MMP26* result, we noted a nearby peak in the *HBB* gene, the lead variant of which fell slightly under GWAS statistical significance (rs73402608; p= 9.38 x10^-8^). Given that the rs334 variant in *HBB* is associated with SCD and was not present in the GWAS because it failed the Hardy Weinberg Equilibrium threshold for quality control, we conducted a post-hoc analysis to determine whether the *MMP26* results remained significant after accounting for this variant. We found that rs334 was strongly associated with morphine intolerance (p= 2.03 x10^-21^), although these results were not significant when homozygous individuals were removed (p=0.14785). While the lead *MMP26* variant and rs334 were not in strong linkage disequilibrium (r2= 0.37), when we adjusted for rs334 homozygous individuals, the *MMP26* signal was abrogated (p= 0.02532), indicating that the *MMP26* signal was likely driven by individuals with SCD.

### Joint assessment of social and genetic factors on glycemic control in diabetes patients

We explored the effect of genetic and social factors on measures of glycemic control among patients with type 2 diabetes (T2DM): HbA1c (primary outcome) and glucose (secondary outcome) laboratory values measured after the first T2DM diagnostic code. There were 5,661 individuals with majority African ancestry meeting criteria for inclusion in the diabetes cohort with quality-controlled laboratory measurements drawn after the first documented diabetes diagnosis in the EHR (4,459 for HbA1c; 5,631 for glucose, with 4,429 overlapping between the two analyses), as well as genotyping for *G6PD*, which, as described above, is associated with diabetes complications. Under an X-linked recessive genetic model, the G6PD deficiency trait was significantly associated with lower first post-diagnosis HbA1c levels (beta=-0.20%, p=0.012) and lower median HbA1c levels within one year of the first measurement (beta=-0.20%, p=0.005). G6PD deficiency trait was therefore included as a covariate in downstream analyses.

G6PD deficiency trait was not significantly associated with first or median post-diagnosis glucose levels in the one year following the first measurement (p>0.05).

To explore the value of integrating genetic and social factors in models of glycemic control, we first sought to identify a single genetic predictor explaining the maximal amount of laboratory trait variance. Polygenic risk scores (PRS) with SNP weights derived from multiple discovery GWAS (**Table S6**) were tested for association with each laboratory trait (**Table S7**). The PRS with the highest proportion of African-ancestry individuals and the strongest association with laboratory traits was derived from GWAS of T2DM in 432,648 individuals of European ancestry and 116,014 individuals of African ancestry in the Million Veteran Program and was selected for use in subsequent analyses^38^. In linear regressions adjusting for age, sex, G6PD deficiency trait, presence of diabetes medications, and 10 PCs for ancestry, the selected T2DM PRS explained more variance in the median post-diagnosis lab values than the first post-diagnosis values (median HbA1c, adjusted R^2^=3.8%, first HbA1c, adjusted R^2^=3.4%; median glucose, adjusted R^2^=2.0%; first glucose, adjusted R^2^=1.4%; **Table S7**). Median post-diagnosis lab values were carried forward for further analysis.

Next, we conducted analyses to determine the contributions of PRS, social factors, and their interactions to variation in measures of glycemic control in T2DM. We chose to focus on financial insecurity, which has been previously established as a risk factor for worse diabetes outcomes^39,40^. As previous EHR-based phenotyping work has demonstrated the value of leveraging both structured and unstructured data sources^41^, we focused on available indices of financial insecurity from both census tract-level data from the American Community Survey (ACS) and data extracted from clinical notes using previously described natural language processing (NLP) methods^42^. Across both laboratory traits and financial insecurity indices, multivariate linear regression demonstrated that T2DM PRS and financial insecurity contributed additively to laboratory trait variation (**Figure 5A**). PRS and financial insecurity indices were both significant and independent predictors of glycemic control, yet the joint model only explained marginally more variance (<1%) than clinical and genetic covariates alone (sex, age, G6PD deficiency, medication status, PCs 1-10). We additionally identified a nominally significant effect of the interaction of T2DM PRS and ACS financial insecurity on median glucose levels (interaction term p=0.047). Differences in T2DM PRS effect size were most pronounced between the top and bottom quartiles (Q1 and Q4) for ACS financial insecurity (**Figure 5B**, Q1, beta= -0.56 mg/dL per SD unit increase in PRS, p=0.67; Q4, beta=5.79 mg/dL per SD unit increase in PRS, p=8.02e-05), indicating that genetic effects of PRS on glycemic control could be more pronounced at increased levels of financial stress. No associations between T2DM PRS and financial insecurity indices were observed (p>0.05), indicating a low suspicion for interaction effects being driven by gene-environment correlation.

**Figure 5:**
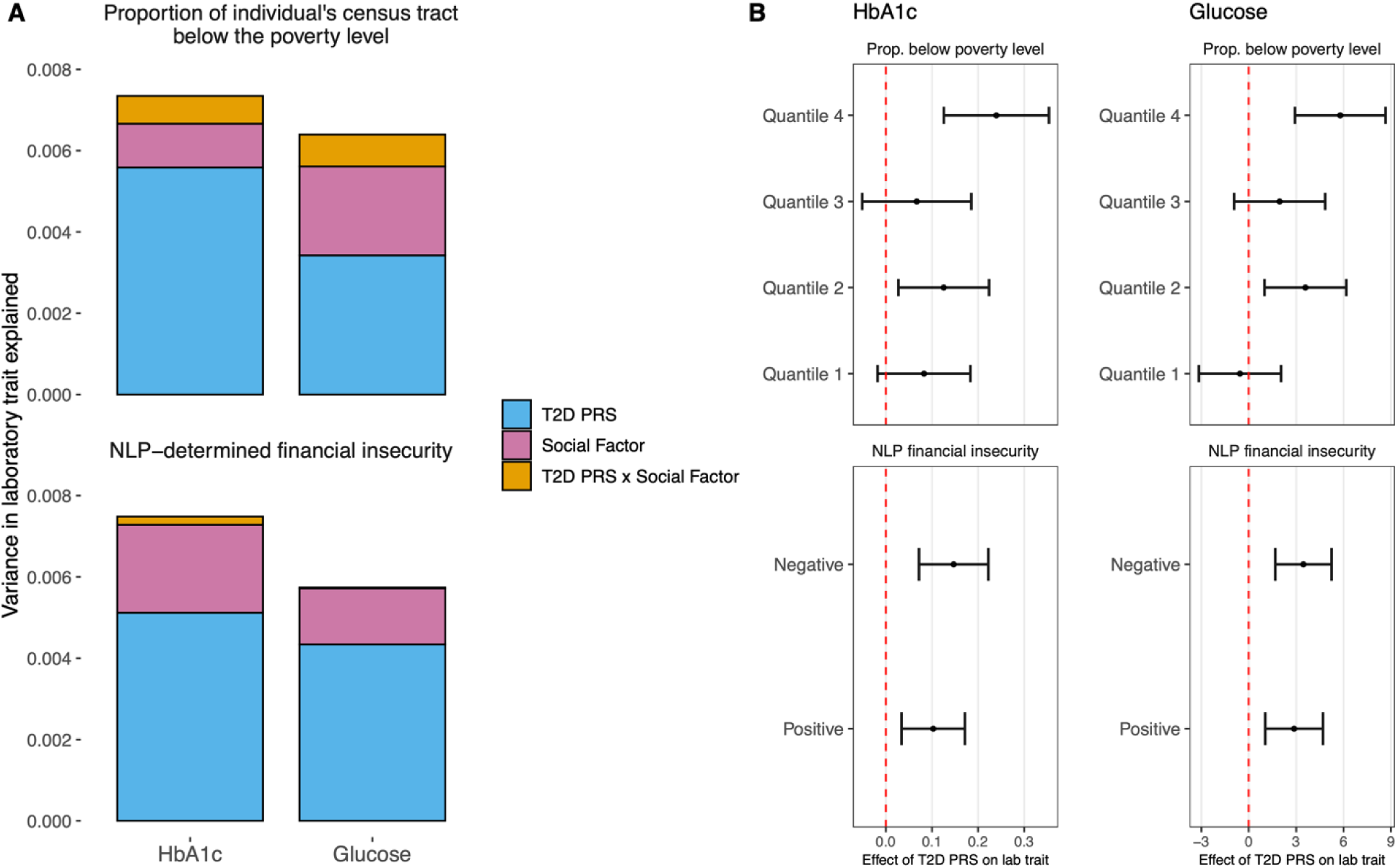
Evaluation of the joint influences of type 2 diabetes polygenic risk scores (T2D PRS) and financial insecurity indices on glycemic control in T2D. **A)** Estimated variance in laboratory trait explained (R^2^) by T2D PRS (blue), social factors (pink), and their interaction (goldenrod) in three separate linear regressions adjusting for clinical and genetic covariates: T2D PRS alone; T2D PRS and social factors combined, and the full model with a term for the interaction between T2D PRS and social factors (T2D PRS x social factors). For visualization purposes, the variance explained due to clinical and genetic covariates is not shown. **B)** Estimated effect of PRS on each lab trait (95% confidence intervals), within strata of financial insecurity indices. Abbreviations: T2D, Type 2 Diabetes; PRS, polygenic risk score; HbA1c, hemoglobin A1c; NLP, natural language processing.

## Discussion

Large, deeply phenotyped cohorts linking EHR data with whole-genome sequencing can advance our understanding of disease and improve clinical outcomes. Including individuals of African ancestry enhances the discovery of genetic associations. Findings from the AGD35k cohort reveal under-documentation of treatable conditions, uncover novel variants, and emphasize the value of social factors in risk assessment. This work highlights the importance of studying genetic variation present across ancestral populations in advancing research and improving healthcare.

Nearly 80% of AGD35k participants had majority African ancestry which is not necessarily indicative of how a person might identify or be identified in the EHR. Although EHR-reported race and ethnicity were used as a rough proxy to enrich the cohort for African genetic ancestry, we focused on ancestry in our analyses as the former are social constructs, have changed throughout time and geographic space, and are insufficient categories to parse human biological variation^43^. Furthermore, due to historical processes related to colonialism, many people, and especially people from socially minoritized populations, have ancestries from multiple geographic regions and often are reported as “Other” or “Unknown” which was frequently the case in our cohort^44^. It is also important to note that over-reliance on continental (global) ancestry can potentially obscure the true scope of an individual’s ancestry and run the risk of excluding important components of variation as seen in the example of pathogenic variants of *TTR* seen in majority European ancestry individuals. This level of fine-grain ancestral inference, that is responsive to the mosaic nature of ancestry across the genome, can improve our understanding of an individual’s disease risk and place it into context of their genetic ancestry.

Clinical genetic testing remains inconsistent, incomplete, and inequitable, leading to missed opportunities for earlier diagnosis and improved outcomes. Despite the potential for severe illness, many carriers of pathogenic *APOL1* and *TTR* variants lacked clinical documentation of a diagnosis even when concordant symptoms were present. In contrast, sickle cell trait, a largely benign genotype, was more frequently documented due to the decades of newborn screening^45^ and strong public health advocacy efforts since the 1970s that has led to potentially curative therapies^46^. However, sickle cell is an exception among pathogenic genotypes primarily affecting patients of African descent. This disparity is especially critical for hATTR, where early intervention with new treatments can significantly improve outcomes. Under-recognition may stem from late onset, heterogenous presentation, variable penetrance or provider unfamiliarity.

Patients with more severe presentations were more likely to raise clinical suspicion and be evaluated for hATTR. Further, individuals of African descent are less likely to receive genetic testing, potentially contributing to lower recognition^47–51^. Increased surveillance for the most frequent hATTR risk variant (V142I) could improve patient care in this population^52^.

We observed no association between sickle cell trait and increased risk for any condition, in contrast to a previous study of individuals with African ancestry in UK Biobank^53^. However, G6PD deficiency was associated with a decreased risk of prediabetes, potentially related to underestimation of glycated hemoglobin and the resulting delayed recognition of diabetes, as recently noted^4^. Both genotypes exhibited distinct sub-clinical differences in RBC parameters typically measured in complete blood counts. Other findings, such as hyposthenuria in sickle cell trait and mild increases in hemolytic markers in G6PD deficiency, suggested physiological differences that were not severe enough to lead to an increased risk of disease. Given the under-documentation of these genotypes, these subtle differences in commonly measured labs could be utilized to prioritize genetic screening for patients. In a recent study, an algorithm using only three laboratory measurements—RBC distribution width, blood glucose, and HbA1c—was developed to predict G6PD deficiency status due to rs1050828 in Black patients and showed promise in identifying individuals for further testing^54^. These results demonstrate the potential for lab-wide scans of common biomarkers to identify subclinical physiologic changes and inform precision medicine strategies.

We observed different ADE frequencies between participants with majority African ancestry compared to those previously reported for individuals with majority European ancestry. For example, ADEs associated with ACE inhibitors occur at a rate of 6.4% in African ancestry participants, while in European ancestry individuals, the rate is 2.4%^35^. This observation is consistent with previous reports that Black individuals are at 3-fold or more risk of serious ACE inhibitor related adverse events compared to White individuals^30,55,56^. We identified 7 loci-drug ADE pairs including an association between rs73407039 in *MMP26* and morphine ADE that is consistent with higher frequency of morphine use among individuals with sickle cell disease. Additionally, we identified novel variant associations to disease that were only possible due to the higher allele frequency in this population. However, when allele frequencies were similar in other ancestral populations, we identified similar association effect sizes. We did not see differences in age or gene-driver distribution of CHIP between African and European ancestries, consistent with the understood stochastic nature of the biological process.

Social and behavioral factors can be extracted from narrative clinic notes in the EHR^42^ and derived from EHR-linked geocoded variables using community surveys ^57,58^. We combined demographic, genetic and social factors to test for association with glycemic control among patients with T2DM. Socioeconomic status is significantly tied to diabetes risk, but it is unclear how it combines or interacts with other risk factors. We show that both genetic and social factors contribute, with slightly larger contributions from genetics, and with some evidence of interaction, to worse glycemic control. As such, only when thinking more holistically can we appreciate the overall picture of risk. Prior work has revealed complex relationships among financial insecurity, structural racism, and health outcomes^59^. While we are unable to assess these associations with the currently available data, our findings should be considered in this context.

Despite the depth and power of this sample, there are several limitations. Participants represent only a small fraction of genetic variation that exists across the African continent. Sample sizes for specific conditions remain smaller than corresponding European samples requiring pooled analysis across the many biobanks with African ancestry participants to power comparable studies. Though EHRs weren’t designed for research, they’ve proven valuable for genomic studies using simple phenotypes like diagnostic codes, and their utility is likely to grow with tools like AI to capture more complex phenotypes. Here, we use multiple different phenotyping approaches including NLP to extract social factors to demonstrate the depth of phenotyping possibilities and richness of the resource.

In summary, we present a series of genetic and phenotypic analyses on a predominantly African ancestry sample demonstrating the power of EHR-linked biobanks such as AGD35k and highlighting important areas of insight for biological and clinical understanding.

## Methods

### Sample selection

For the selection of subjects for AGD35k, the BioVU^®^ population meeting DNA sample eligibility (n=287,354; based on data from the December 2022 version of the Synthetic Derivative [SD], i.e., data through October 31st, 2022) was first bifurcated based on MEGA^EX^ genotyping status. Among those already genotyped (n=83,990), 13,371 subjects with known relative majority African ancestry were selected. Among those not selected, propensity scores were used to predict relative majority African ancestry based on EHR-reported race and ethnicity. These scores were calculated by first splitting the total MEGA^EX^ population (excluding those without separate first/last EHR dates based on full scope of OMOP data availability, such as ICD/CPT codes, notes, medications, etc.) into training and testing sets before modeling on the training set with logistic regression (n=87,903). Following confirmation of score > 0.02 in the test set with a sensitivity of 0.98 and specificity of 0.27, scores were then predicted for all remaining subjects not already selected for known relative majority African ancestry, first based on a threshold of score > 0.1 (n=22,051), followed by a random selection of subjects with score > 0.08 (n=968), in order to meet the target number of total subjects (n=23,019; n=36,750 total).

### Sample Preparation for WGS

Samples were prepared for sequencing at deCODE Genetics using the Illumina DNA PCR-Free Prep (IDPF) method, product # 20041795 (https://www.illumina.com/products/by-type/sequencing-kits/library-prep-kits/dna-pcr-free-prep.html). The method entails the use of a bead-linked transposome (BLT), in which an enzymatic reaction referred to as “tagmentation” is used to both fragment genomic DNA and ligate sequencing adaptors to the resulting fragments in a single step. The beads serve to normalize the amount of sequencing libraries generated for each sample.

In short, approx. 500 ng of genomic DNA (25 µL at 20 ng/µL) was used as input in a 96-well plate format. DNA samples were quantified using the Lunatic UV/Vis next-gen plate reader (https://www.unchainedlabs.com/lunatic/) and diluted appropriately. Sample preparation was performed using a fully automated workflow on Hamilton STAR NGS liquid handlers (96- channel pipetting) with validated scripts from Illumina. Processing of each plate was completed in approx. 90 min. Samples were indexed with 2*10 base molecular barcodes using one of four available dual index sets (A-D), for a maximum of 384 indexes (IDT for Illumina DNA/RNA UD Indexes, products #20027213, #20027214, #20042666, #2004267). Sequencing libraries were stored at -20 °C. until use. All steps in the workflow were recorded using an in-house laboratory information management system (LIMS) with barcode tracking of all samples and reagents.

### Sequencing and variant calling

At least two rounds of sequencing were performed at deCODE Genetics for each sample. In the first round of sequencing, each library plate was pooled in equal volume per library (5 µL). The pools were quantified using the Qubit™ ssDNA BR assay kit (Thermo-Fisher), diluted appropriately in resuspension buffer (RSB), denatured, and stored frozen at -20 °C until sequencing, All steps in these procedures were done using Hamilton STARlet liquid handlers (8- channel pipetting). Library pools were sequenced on Illuminás NovaSeq 6000 instruments, using S4 flowcells and the SBS v.1.5 sequencing kits (product #20028312). Paired-end sequencing was performed with run configuration of 151x10x10x151 cycles of incorporation and imaging using the standard sequencing workflow, i.e,. one pool across all four lanes on the flowcell. Basecalling was done in real-time on each sequencer using NovaSeq Control Software version 1.8.0. Downstream processing of sequence data was done using an in-house pipeline which included FASTQ generation (BCLConvert) and alignment to the GRCh38 genome build using the DRAGEN server for the generation of lane .cram files. Quality control of each lane cram file was done using in-house scripts, assessing various sequencing attributes, including GC fraction, base conversions in trinucleotide context, library insert size distribution, and properly paired reads. A second round of pooling for each plate was done based on the average genome coverage for each library. This step was performed in order to normalize the genome coverage of each library across each plate. The volumes were calculated using the in-house LIMS and each library was pooled appropriately, as described above. Each pool contained anywhere from 32 to 48 libraries and was quantified, denatured, and sequenced as described above. Each library was sequenced to an average genome coverage of ≥28X, with ≥95% of all samples across the cohort with coverage ≥30X. All libraries also had at least 15 reads covering 95% of the autosome.

Libraries that failed to reach these criteria following two rounds of sequencing underwent top-up sequencing using the NovaSeq-XP workflow, i.e. individual lane loading (product #20043131) until the sequencing requirements above were fulfilled. Merging of individual cram files was done using Samtools merge (v1.9). Summary statistics were calculated for each merged sample and uploaded to Illumina Connected Analytics (ICA) along with the merged cram files.

Reads from loss-less CRAM files were aligned with Illumina’s DRAGEN version 3.7.8 Germline pipeline to the GRCh38 human reference genome with ALT contigs plus additional decoy contigs and HLA genes (as recommended for GRCh38 mapping). Variant calling on the WGS Data was performed by Illumina using DRAGEN version 3.7.8 in Illumina Connected Analytics. QC was performed, evaluating sequencing yield, genomic coverage, proper-pair mapping, contamination metrics, and genetic sex concordance. Multisample VCFs were created with the DRAGEN Iterative gVCF Genotyper (IGG) pipeline (v4.2.7).

### Variant and sample quality control (QC)

Of the 35,014 eligible participants in AGD35k, 15,313 (44%) also had genotyping on the MEGA^EX^ platform. To assess for sample swaps (i.e., mismatch between an individual’s AGD and MEGA^EX^ genotypes), we identified a subset of 744,305 SNPs present in both datasets and meeting the following criteria: bi-allelic, common (MAF>1%), low missingness (<1%), and non-palindromic. PLINK’s pgen-diff was used to identify SNPs with discordant results on the two platforms. Four samples were discordant representing likely sample swaps and removed from the analysis cohort.

As an additional measure to detect sample swaps, EHR-coded gender was compared to the genetic sex as calculated by Illumina’s DRAGEN ploidy estimator which estimates a sample’s karyotype "using the ratios of the median sex chromosome coverages to the median autosomal coverage". In the AGD35k dataset, 129 participants had discordant genetic sex and EHR-coded gender. No participants labeled as sex discordant were genotype concordance outliers. Therefore, samples were flagged for sex discordance but not removed on these criteria alone.

COMPADRE was used to produce relatedness estimates between individuals in the cohort. Variants with a frequency < 0.05, missingness > 0.10, or ancestry informative were excluded from the analysis. COMPADRE identified 1,075 1^st^ degree pairs, corresponding to 1,892 individuals. This included 3 pairs of monozygotic twins. EHR-reported gender, date of birth, and clinical notes were used to validate pairs of twins versus potential sample duplications. Pedigrees were detected using default settings in COMPADRE. We initially used a relatedness cutoff of 3^rd^ degree to determine pedigrees. If any pedigrees failed to reconstruct, then we iteratively repeated the pedigree reconstruction step for the specific pedigrees with a relatedness cutoff of 2^nd^ and then 1^st^ degree.

Contamination estimates were produced by Illumina’s DRAGEN pipeline and suggested very low contamination across the dataset (< 1%). Estimates of excess heterozygosity and the heterozygous:alt-homozygous genotype ratios, produced by PLINK 2.0, across individuals additionally supported the conclusion that contamination is low. The quality of the sequenced samples was investigated by calculating Ts/Tv ratios. Individuals > 4 standard deviations (SD) from the mean were removed from the cohort (n=31). The median coverage across the dataset is 32.5x. Individuals with less than 90% of their genomes sequenced to 10x coverage were excluded from the cohort (n=17).

Variants flagged by DRAGEN^60^ during variant calling for low quality (e.g., NO_HQ_GENOTYPES, ExcessHet, LowQual) with a filter field ≠ ’PASS’ were excluded. For the remaining variants, individual genotypes were retained if they met the following criteria: genotype quality (GQ) ≥ 20, depth of coverage (DP) ≥ 10, allele balance (AB) between 0.2 and 0.8 for heterozygotes, and a filter field == ’PASS’. Variants with a missing rate below 0.10 and loci with 2-6 alleles were retained.

AGD35k was phased after removing all multiallelic sites and requiring MAF > 0.01 using SHAPEIT5^61^.

### CHIP detection

Clonal hematopoiesis of indeterminate potential (CHIP) was detected using established methods^62^. In brief, whole genome sequence data was filtered to regions of the genome that are known to harbor CHIP driver mutations^63^ and putative somatic mutations were detected with Mutect2 from GATK and annotated with ANNOVAR^64,65^. Putative somatic mutations were filtered to exclude those at loci with fewer than 15 total reads, fewer than 2 reads from the second most common allele, or an absence of the second most common allele in either sequencing direction. Hotspots were defined as genomic loci with more than 15 putative somatic variants within the sequenced population and were excluded as likely sequencing artifacts if the putative somatic variant lacked association with both age and the *TERT* promoter variant known to drive CHIP (rs7705526) defined by *p* > 0.1 in logistic regression analysis.

### Principal Component Analysis

We used the 1000 Genomes Project phase 3 (1kGP, high coverage dataset as provided on the PLINK resource page) for performing principal component analyses^66^. Data were filtered to remove 629 related individuals (KING cutoff 0.0884, 2,573 samples remaining). The unrelated 1kGP set was merged with AGD35k (n=35,024) using the intersection of variants (merged 1kGP+AGD35k set). To create principal components (PCs) of genetic ancestry, we projected PCs using the Human Genome Diversity Project and 1kGP allele frequency and loading files provided by the Global Biobank Meta-analysis Initiative (GBMI)^67^. The merged 1kGP+AGD35k set (n=37,597) was filtered to the GBMI variant set (140,535 variants). Samples were then projected into the same PC space with the plink2 –score option, extracting 20 PCs^68^.

### Global Ancestry Estimation

We inferred admixture proportions in the 1kGP+AGD35k set using SCOPE, which estimates an individual allele frequency matrix through latent subspace estimation and then decomposes the estimated matrix into ancestral allele frequencies and admixture proportions^69^. The 1kGP+AGD set was filtered to remove rare and multiallelic variants (MAF < 0.01). Long-range linkage disequilibrium (LD) regions within GRCh build 38 were filtered^70^, before a final LD filter was applied ("--indep-pairwise 50000 80 0.1") resulting in 399,846 remaining variants. Unsupervised SCOPE was run, setting the number of inferred populations k=5. The SCOPE supervised mode uses allele frequencies in a reference population. Within the 1kGP dataset, we filtered out duplicate, multiallelic, and rare variants (MAF < 0.05), and calculated the allelic frequencies within 1kGP superpopulations (AFR, AMR, EAS, EUR, and SAS) using PLINK2. The filtered 1kGP+AGD 25K set was restricted to 122,521 variants and then supervised SCOPE was run. For both supervised and unsupervised analyses, individuals were assigned to the ancestry cluster for which the proportion exceeded 0.5; individuals without an ancestry proportion exceeding 0.5 were assigned as “Admixed – majority ancestry proportion < 0.5”.

### Local Ancestry Inference

In preparation for determining local ancestry in AGD35k, we selected the Koenig et al., 2024 reference population to best capture the genetic diversity of our dataset, as it is a harmonized reference population spanning the Genome Aggregation Database (gnomAD), 1000 Genomes Project (1kGP), and Human Genome Diversity Project (HGDP) with seven super-populations (African [AFR], European [EUR], Central/South Asian [CSA], East Asian [EAS], Oceanian [OCE], Middle Eastern [MID], Admixed American [AMR])^71^. The reference population was filtered to remove admixed individuals (from super-populations and subpopulations, 193 individuals removed) and related individuals (3 individuals removed), to ensure accurate local ancestry inference. The remaining reference population (n=3,407) was filtered to remove rare variants (MAF > 0.01) for each individual superpopulation, and the resulting data was phased using SHAPEIT5^61^. Using our phased reference population and phased AGD35k data, local ancestry of AGD35k was inferred using RFMIX2^72^. Global ancestry proportions were also generated by RFMIX2 and resulted in highly concordant results with the global ancestry estimates from the SCOPE analysis (r^2^ > 0.8, within individual Pearson’s correlation coefficient (r^2^) were calculated to check for consistency of ancestry proportion inferences between the two methods).

To evaluate the local ancestry at the haplotypes containing the TTR amyloidosis pathogenic variants, we extracted the ancestral population assignments at chromosome 18, starting position 31555042 and ending position 31731379 (GRCh38) from the RFMIX2 results. This haplotype contains all the observed TTR pathogenic variants identified within the AGD35k population (TTR c.130C>T, TTR c.148G>A, TTR c.238A>G, TTR c.239C>T, TTR c.379A>G, TTR c.424G>A). We restricted our local ancestry analyses to the more common V142I (n=1,102). We calculated the number of haplotypes mapped to each genetic ancestry group at the TTR locus (AFR/African, CSA/Central South Asian, EAS/East Asian, EUR/European, MID/Middle Eastern, or OCE/Oceania) in all AGD35k individuals (including TTR carriers and non-carriers (n=1,102 and n=33,916, respectively)). We then performed a logistic regression model to determine if African ancestry at this locus was associated with TTR carrier status. Additionally, we used a linear regression model to test whether global African ancestry proportion was associated with TTR carrier status. We used the AncestryGrapher toolkit Python package^73^ to generate the painted chromosome 18 karyograms.

### Phecode Dataset

Phecode files were generated for patients in the AGD35k dataset with at least 3 unique visit dates 30 days apart for inclusion in the phenome cohort. We mapped ICD9CM and ICD10CM codes to phecodes for both phecode 1.2 and phecode X definitions, using the phecode 1.2 to ICDCM unrolled map and the phecode X to ICDCM unrolled map, respectively, sourced from the phewascatalog.org site. These phecodes were translated from a set of patient ICD codes drawn from the Vanderbilt Synthetic Derivative (SD) using the: CONDITION_OCCURRENCE, OBSERVATION, MEASUREMENT, PROCEDURE_OCCURRENCE, and DRUG_EXPOSURE tables as defined in the OMOP common data model. Only billing codes were included, excluding problem list codes.

Analysis

### CHIP PheWAS

For PheWAS of CHIP status, the standard PheWAS approach was inappropriate, due to the time-varying nature of CHIP^74^. We applied a survival-PheWAS approach using a Cox proportional hazards model where time at risk started at the date of blood sample collection, events were defined by first occurrence of a given phecode, and right-censoring was determined by date of last encounter with the medical system. Covariates included age at blood sample, age squared, genetic sex, EHR-coded race, and smoking status. Research participants whose date of first diagnosis with a given phecode preceded sample collection date (prevalent disease) were excluded from survival analysis.

### GWAS of phecodes and comparison across ancestries

We retained biallelic variants with a missing rate < 0.02 and a MAC ≥ 100 from AGD35k for GWAS. Variants on autosomes and in the pseudoautosomal region (PAR) of chrX were further filtered using a HWE p-value threshold of < 1×10 ¹ . There were 27,782 AFRs with 17,831,304 QCed variants and 5,130 EURs with 8,282,531 QCed variants for analysis.

To efficiently perform GWAS across thousands of phecodes, we developed a workflow that leveraged Regenie v4.1. In Regenie Step 1, we selected unambiguous SNVs from the EUR and AFR subsets with a missing rate < 0.01 and MAF > 0.05. After removing known high-LD regions, we performed LD pruning using PLINK (--indep-pairwise 1000 50 0.05) to generate a high-quality, independent variant set, which was then used to construct the whole-genome regression model. These variants were divided into consecutive blocks of 1,000, and ridge regression predictions were computed for each block. The predictions were subsequently aggregated using a second ridge regression and partitioned by chromosome to enable leave-one-chromosome-out (LOCO) analysis. In Regenie Step 2, using the same EUR and AFR subsets, we conducted single-variant association testing with Firth logistic regression for the analysis of binary traits (as default setting), conditioning on the Step 1 predictions. For variants with nominal p-values < 0.05, an approximate Saddle Point Approximation (SPA) correction was applied. These two models from step 1 and step 2 included sex, baseline age, age², and the first 20 principal components (PCs) as covariates.

For phecodes with a prevalence greater than 10% in the AFR population, we conducted GWAS separately in both the AFR and EUR ancestry samples using the default settings of the workflow described above. To identify top independent association signals, we applied LD clumping to the GWAS summary statistics using PLINK2 with the parameters --clump-kb 500, --clump-p1 5e-8, --clump-p2 0.005, and --clump-r2 0.1. This procedure identified 199 genome-wide significant loci in AFR and 23 in EUR across 96 phecodes. We removed 79 associations driven by participants with SCD, 10 driven by participants with APOL1 and 8 on the X chromosome where prior association results remain more limited.

Using the 5,879 SNP/phecodes associations in the Phenotype-Genotype Reference Map (PGRM) as a reference, we assessed replication among the 122 GWAS studies conducted in AFR and EUR populations that included variants with a MAF > 0.01. These summary statistics were annotated separately for AFR and EUR ancestries using the annotate_results() function from the pgrm R package. Finally, we calculated the overall replication rate (RR_All) and the powered replication rate (RR_Power) for each ancestry group by applying the get_RR() function to the annotated results.

### Evaluating clinical recognition and expression of ancestry-specific genetic variants

#### Identifying monogenic carriers

We identified carriers of pathogenic variants in the transthyretin (*TTR*) gene, associated with hereditary TTR amyloidosis; the apolipoprotein L1 (*APOL1*) gene, associated with APOL1-mediated kidney disease; the glucose-6-phosphate dehydrogenase (*G6PD*) gene, associated with G6PD deficiency; and the hemoglobin subunit beta (*HBB*) gene, associated with sickle cell disease. Participants were defined as having sickle cell trait or disease if they were carriers of one or two copies of the rs334 (c.20A>C) variant, respectively. Any participant with sickle cell trait who had a secondary, deleterious variant was excluded from the sickle cell trait and disease analyses. This included 183 compound heterozygous participants with a secondary variant in addition to HbS: 134 with an HbC variant, 33 with a β+ variant, 9 with a β0 variant, 3 with an Hb Korle-Bu variant, 2 with an HbE, variant, and 2 with an HbO. Participants were defined as at-risk for G6PD deficiency if they were carriers of two copies of the rs1050828 missense variant (c.292G>A) which is known to cause G6PD deficiency.

Participants were defined as having the risk genotype if they carried one copy of the risk genotype if male or two copies of the risk genotype if female. Participants were defined as at-risk for APOL1-mediated kidney disease if they were carriers of two alleles of either the G1- rs71785313 (c.1024A>G) and rs60910145 (c.1152T>C) variants, or the G2-rs73885319 (g.36266000_36266005del) variant. Participants carrying no risk allele or only one risk allele in *APOL1* were defined as not at risk. Participants were defined as at risk for hereditary TTR amyloidosis if they were carriers of any of 55 selected variants identified in ClinVar (downloaded March 2024). These variants were selected if they were reported as pathogenic or likely pathogenic and two-star review status. Six of the variants queried were detected within our cohort. Participants that did not carry the queried alleles were defined as not at risk.

#### Defining clinical ascertainment

To define clinical recognition of disease risk variants, we utilized a combination of diagnosis codes (ICD/phecodes), testing (CPT/HCPCS) codes, and free-text clinical notes queried by specific keywords. Structured data sources used to define ascertainment are available in **Table S2**.

#### Lab values extraction and curation

We selected 33 laboratory measurements from five laboratory panels that are measured for screening purposes—the complete blood count, the basic metabolic panel, liver panel, hemoglobin A1c/random blood glucose, and urine albumin-creatinine ratio—as these are less likely to be impacted by ascertainment bias^75^. Clinical laboratory measurements were processed according to a previously described standardized bioinformatic pipeline using data from the VUMC SD^76^. Briefly, clinical laboratory lab data was filtered to only observations with positive, numeric values. Additionally, only observations with non-missing reference ranges were kept. For each lab, we ascertained the most common unified codes of units of measurement (UCUM) unit and restricted to observations in this UCUM unit.

#### Association analyses of sickle cell trait and G6PD deficiency

To assess the clinical implications of genotype carrier status, PheWAS was performed using both billing codes (phecodes) and laboratory values as outcomes^76,77^. For each phenotype, logistic regression was used, defining cases as participants who had one or more of a given phecode. For each laboratory test, linear regression was used to test the association between genotype and the inverse-normal transformed median value among the subset of participants with a given lab measurement. All analyses were adjusted for sex, age, age-squared, and 10 PCs. The age for each participant was defined as the median age across all phecodes for the phecode analysis and the median age of the specified laboratory measurement for the laboratory analysis. Analyses were repeated after restricting to participants with majority African ancestry and after excluding participants with a known diagnosis to assess whether under-diagnosis was clinically significant. Because the inverse-normal transformed lab values are not clinically interpretable, the lab analysis was repeated using the unadjusted laboratory values. Multiple testing corrections were applied using a Bonferroni correction. Statistical significance was defined as a p-value of 1.9×10^-4^ (33 distinct labs × 2 genotypes × 4 strata) for the lab analysis and a p-value of 1.7×10^-6^ (3,613 phecodes × 2 genotypes × 4 strata) for the PheWAS analysis.

### TTR Carrier Filtering, Phenotype Characterization, and Chart Review

We selected 55 pathogenic variants in TTR from ClinVar with 2-star review status. We filtered individuals who carried risk alleles from the cohort and separated them by zygosity. TTR-associated phecodes “Heart failure”, “Polyneuropathies”, “Amyloidosis”, “Cardiomyopathy”, and “Mononeuritis of the upper limb” were selected based on literature review and were limited to highly replicated features of hereditary TTR-amyloidosis. We further filtered our analysis to individuals whose age at their last clinic visit was over the age of 30 to account for the age-related penetrance associated with this disease. ICD9 and 10 codes in the records of the remaining participants were mapped to phecodes, and filtered for the presence of any of the five previously defined TTR-associated phecodes. Of note, the “Amyloidosis” phecode maps to ICD- 10 code E85 and ICD-9 code 277. These are not specific to TTR amyloidosis, and the context of their use was a central goal of downstream chart review. We also assessed the frequency of these phecodes in non-genetic risk carriers to calculate the population attributable fraction of these symptoms due to TTR genetic risk. CPT codes relevant to TTR work-up and diagnosis, such as EKG, cardiac MRI, Congo red staining, and 99mTC-PYP were also extracted from the EHR for all genetic risk carriers. The frequency of these codes in genetic risk carriers versus non-genetic risk carriers were assessed in a chi-squared test.

Individuals who had at least one relevant phecode in their record were selected for manual chart review. Chart review chiefly aimed to identify which individuals received a diagnosis. We also made note of what level of specificity (i.e. genetic testing) or clinical test. Clinical notes were used to determine if family history was discussed, if TTR amyloidosis was ever considered as a differential diagnosis, and first mention of amyloidosis (if present).

### Joint assessment of social and genetic factors on glycemic control in participants with diabetes

#### Diabetes cohort

Diabetes was defined as having two outpatient diabetes codes on different dates, one diabetes code, and a qualifying HbA1c > 6.5, or having a diabetes code and being on diabetes medication. Participants with gestational diabetes or only one diabetes code were excluded. The date of the first diabetes code was considered the date of diabetes diagnosis and was required to occur at age 18 or older. The primary outcome was HbA1c, and the secondary outcome was blood glucose.

#### Extraction and curation of social factors from EHR

##### ACS-derived financial insecurity

Structured indices of financial insecurity were derived from responses to the American Community Survey (ACS) aggregated at the level of census tracts. Specifically, the percentage of census tract below the poverty level was used. Census tract-level poverty data were de-identified to adhere to HIPAA Safe Harbor Guidelines using a constraint-based *k*-means model to group similar tracts together such that each cluster has >20,000 inhabitants.

##### NLP-derived financial insecurity metric

We also employed our previously developed Phenotype Retrieval system, PheRe^41,42,78,79^, to identify participants experiencing financial insecurity. We have proven the utility of PheRe in multiple diverse use-cases, including social factors, suicidality, sleep-related behaviors, and more^42,78–80^. Briefly, PheRe is a search engine designed on top of an information retrieval architecture able to process a large volume of clinical notes for a given query describing a phenotype—financial insecurity, in this case). The input queries describing the phenotypic profile of the NLP feature were constructed leveraging data-driven and domain expertise methodologies. The data-driven methodology involved 1) processing large collections of clinical notes to learn context-independent and context-sensitive word embeddings; 2) iteratively expanding an initial set of high-relevant expressions (also called ‘seeds’); 3) ranking the learned embeddings by their similarity to the seed embeddings; and 4) manually reviewing the top ranked expressions (i.e., in the embedding space) for selecting the relevant ones as new seed expressions. The financial insecurity queries used in this study are listed in **Table S8**. We modeled financial insecurity as a binary variable, with “positive” indicating the presence of any query term in the patient’s notes.

#### Diabetes related variable extraction and QC

##### Laboratory tests

Glucose and HbA1c laboratory values (OMOP concepts 3004501, 3000483, 3005673, and 3004410) for individuals in the diabetes cohort were extracted from the Synthetic Derivative, and units were harmonized using the UCUM unit library. Implausible glucose (<40 mg/dl or >400 mg/dl) and HbA1c (<2.5% or >25%) values were removed. Next, values four SD above the cohort mean were removed for each lab. Lab measurements were restricted to those drawn after the first diabetes diagnosis code. Two variables were defined and used as outcomes in downstream analyses: 1) the first lab drawn after the first diagnosis, and 2) the median lab value in the year following the first lab drawn after the first diagnosis.

##### Medications

Medications were captured from OMOP tables using drug concept mapping, primarily drug concept names. Drug captures included all current diabetic drugs available (insulin, sodium-glucose cotransporter-2 inhibitors (SGLT2i), Glucagon-like peptide-1 receptor agonists (GLP-1 RAs), metformin, sulfonylureas, Dipeptidyl peptidase 4 (DPP-4) inhibitors DPP4 inhibitors, alpha-glucosidase, meglitinide analog, amylin analog, and thiazolidinediones. For regression analyses, individuals were classified as being on diabetes medication if there was evidence of any of the queried medications within one year prior to the first post-diagnosis lab measurement.

##### Polygenic risk scores

Using the common variant QC-ed version of AGD35k described above, SNVs were mapped from Genome Reference Consortium Human Build 38 to Build 37 coordinates. To ensure high-quality SNPs, the SNVs were selected from the HapMap3 subset data, resulting in 27,804 participants with majority African ancestry (5,142 of EUR ancestry) and 1,244,670 common SNVs (1,215,651 for EUR ancestry) for analysis. We downloaded the African and European ancestry linkage disequilibrium reference panel from the 1000 Genomes Project Phase 3 and kept non-ambiguous SNPs with MAF > 0.01, using the largest European ancestry GWAS summary statistics for BMI, T2D, and HbA1c (references listed in **Table S6**). We manually harmonized strand-flipping SNPs among the SNP information file, GWAS summary statistics files, and the African ancestry PLINK extended map files (.bim). We used PRS-Continuous Shrinkage (PRS-CS)^81^ to infer posterior SNP effect sizes under continuous shrinkage priors with a scaling parameter of 0.001. We additionally tested scores generated using PRS-CSx (PMID: 35513724) with the “meta” flag, leveraging cross-population discovery GWAS and LD panels constructed from both European and African reference populations, using a scaling parameter of 0.01. The scoring command in PLINK version 1.9 was used to produce genome-wide polygenic risk scores (PRS) for the African- and European-ancestry participants based on their quality-controlled SNP genotype data and the derived SNP weights. The “MVP_CSx_META_t2d” scores, which had the highest genetic similarity between the training and target samples (i.e., the largest number of African-ancestry individuals in the training GWAS) and was most strongly associated with the laboratory outcome variables in the diabetes cohort, were retained for analysis (**Table S7**).

#### Integrating social and genetic factors

We conducted an association analysis, among individuals in the majority African ancestry cohort, assessing effect sizes of these multimodal covariates on glycemic control as measured by HbA1c (primary outcome) and glucose (secondary outcome). Given the impact of G6PD deficiency on HbA1c and G6PD deficiency prevalence in those of AFR ancestry, we also included the *G6PD* genetic variant as defined above in these analyses.

To emphasize interpretability of associations, we used non-regularized linear regression with HbA1c and glucose as the dependent variables and the clinical (presence of diabetes medications), genetic (*G6PD* genotyping, T2D PRS), and financial insecurity (structured and unstructured) features as independent variables. We accounted for interactions between polygenic scores and financial insecurity by specifying them manually. Covariates for age at first post-diagnosis lab draw, genetically determined sex, diabetes medication status, G6PD deficiency trait, and 10 PCs for ancestry were included in all regression analyses. ACS-derived financial insecurity was modeled continuously in these analyses. A post-hoc test was conducted to identify the effect of T2D PRS on glucose levels between individuals in the top and bottom quartiles for ACS-derived financial insecurity.

We evaluated model goodness-of-fit with R-squared for models comprised of subsets of features by category: clinical and genetic covariates only, polygenic scores and covariates, financial insecurity and covariates, full model with all features, and full model plus a term for the interaction between polygenic score and financial insecurity. Further, for each regression model, we estimated relative contributions of T2D PRS, social factors, and their interaction by calculating R^2^ values for a series of nested regression models where each term was added iteratively.

### Pharmacogenomics

#### High-throughput Identification of Adverse Drug Effects (ADE) in EHRs

We identified ADEs using a modified version of a previously published high-throughput natural language processing approach using regular expressions^82^. We targeted 11 frequently prescribed drugs/drug classes with common adverse events: ace inhibitors, cephalexin, CYP2D6 mediated opioids, erythromycin, levofloxacin, meperidine, morphine, penicillin, statins, sulfas, and aspirin.

Medications were identified in the allergy section of the clinical notes by searching for case-insensitive regular expressions for generic names, brand names, and abbreviations (e.g., "pcn" for penicillin). A random sample of notes (n≥1000) for each drug/drug class was reviewed by experts to assess the need for any additional expressions or revisions for systemic false positives to avoid misclassification. If the list was revised, we completed additional reviews (n≥1000), iterating as needed until systemic issues were addressed (max iterations n=4). **Table S4** includes a complete list of the regular expressions used to identify the 11 drugs/drug classes. For each drug/drug class, we defined cases as individuals with any mention of the drug in the allergy section and controls were individuals with no known drug allergy (i.e., no mention of the corresponding drug in the problem list of their EHR).

During review, we noted that aspirin was included in the allergy list due to comorbidities that preclude its use (approximately 3.5%). As such, we excluded patients with notes indicating comorbidities with noted contraindications from the analysis, including "kidney," "renal," "sickle cell," "Willebrand," "hemophilia," "hepatitis," " PUD," "ulcer," “Crohn,” and "contraindicated" (**Table S4**). We also excluded brand names for drug combinations that included aspirin (e.g., “Excedrin”), as well as entries that included medications that are frequently combined with aspirin which could be the source of the ADE (e.g., "hydrocodone," and "codeine") to limit misclassification. These patterns of potential misclassification were not evident in the notes for other drugs/drug classes.

#### Statistical Analysis

We used the same GWAS pipeline described above to determine whether there was an association between the adverse drug effect and common genetic variants (i.e., n≥200 minor allele count within the ADE cohort), with adjustment for age at most recent clinical visit, sex, EHR length, and 10 PCs for ancestry. We considered p-values less than 5x10^-8^ as significant.

## Data availability

This study used data from the Alliance for Genomic Discovery (AGD), a partnership between Nashville Biosciences, Illumina, and Vanderbilt University Medical Center (VUMC). Data access is subject to restrictions and are made available to commercial researchers via NashBio and academic researchers via BioVU.

Summary statistics of all results including GWAS and PheWAS are provided publicly here https://victr.vumc.org/alliance-for-genomic-discovery/

## Code availability

These data were processed and analyzed using publicly available software as noted throughout the manuscript.

## Supporting information

Complete Author Information

## Acknowledgements

Vanderbilt University Medical Center’s BioVU projects are supported by numerous sources: institutional funding, private agencies, and federal grants. These include NIH funded Shared Instrumentation Grant S10OD017985, S10RR025141, and S10OD025092; CTSA grants UL1TR002243, UL1TR000445, and UL1RR024975. Genomic data are also supported by investigator-led projects that include U01HG004798, R01NS032830, RC2GM092618, P50GM115305, U01HG006378, U19HL065962, R01HD07471; and additional funding sources listed at https://victr.vumc.org/biovu-funding/.

The sequencing of WGS individuals from BioVU, including the 35,024 described here, has been funded by the Alliance for Genomic Discovery consisting of NashBio, Illumina and industry partners Amgen, AbbVie, AstraZeneca, Bayer, BMS, GSK, Merck Sharp & Dohme LLC, and Novo Nordisk. DNA sequencing was performed at deCODE genetics using Illumina sequencing technology.

This work was supported by National Institutes of Health (NIH) grants R01HG012657 (Douglas Ruderfer and Lisa Bastarache), U54CA163072 (Jibril Hirbo), RHL174052A and R01HL159557 (Piper Below), T32 HG008341 (Freida Blostein and Cecile Avery), K12AR084232 (Megan Shuey), 5TL1TR002244 (Jeewoo Kim), Merit award 2I01CX001897-06A1 (Adriana M. Hung), We thank Otis Wilson for aiding clinical data preprocessing. We thank the VANTAGE biobanking team for help with sample processing.

## References

1. Campbell, M. C. & Tishkoff, S. A. AFRICAN GENETIC DIVERSITY: Implications for Human Demographic History, Modern Human Origins, and Complex Disease Mapping. Annu. Rev. Genomics Hum. Genet. 9, 403–433 (2008).

2. Gomez, F., Hirbo, J. & Tishkoff, S. A. Genetic Variation and Adaptation in Africa: Implications for Human Evolution and Disease. Cold Spring Harb. Perspect. Biol. 6, a008524 (2014).

3. Cohen, J. et al. Low LDL cholesterol in individuals of African descent resulting from frequent nonsense mutations in PCSK9. Nat. Genet. 37, 161–165 (2005).

4. Breeyear, J. H. et al. Adaptive selection at G6PD and disparities in diabetes complications. Nat. Med. 30, 2480–2488 (2024).

5. Friedman, D. J. & Pollak, M. R. Genetics of kidney failure and the evolving story of APOL1. J. Clin. Invest. 121, 3367–3374 (2011).

6. Mulder, N. et al. H3Africa: current perspectives. Pharmacogenomics Pers. Med. 11, 59–66 (2018).

7. Abul-Husn, N. S. & Kenny, E. E. Personalized Medicine and the Power of Electronic Health Records. Cell 177, 58–69 (2019).

8. Bycroft, C. et al. The UK Biobank resource with deep phenotyping and genomic data. Nature 562, 203 (2018).

9. Nagai, A. et al. Overview of the BioBank Japan Project: Study design and profile. J. Epidemiol. 27, S2–S8 (2017).

10. Kurki, M. I. et al. FinnGen provides genetic insights from a well-phenotyped isolated population. Nature 613, 508–518 (2023).

11. Chen, Z. et al. China Kadoorie Biobank of 0.5 million people: survey methods, baseline characteristics and long-term follow-up. Int. J. Epidemiol. 40, 1652–1666 (2011).

12. Bick, A. G. et al. Genomic data in the All of Us Research Program. Nature 627, 340–346 (2024).

13. Gaziano, J. M. et al. Million Veteran Program: A mega-biobank to study genetic influences on health and disease. J. Clin. Epidemiol. 70, 214–223 (2016).

14. Roden, D. et al. Development of a Large-Scale De-Identified DNA Biobank to Enable Personalized Medicine. Clin. Pharmacol. Ther. 84, 362–369 (2008).

15. Hulsizer, J. et al. Sickle Cell Trait and Risk for Common Diseases: Evidence from the UK Biobank. Am. J. Med. 135, e279–e287 (2022).

16. Naik, R. P. & Derebail, V. K. The spectrum of sickle hemoglobin-related nephropathy: from sickle cell disease to sickle trait. Expert Rev. Hematol. 10, 1087–1094 (2017).

17. Khaled, S. A. A. et al. Hematological, Biochemical Properties, and Clinical Correlates of Hemoglobin S Variant Disorder: A New Insight Into Sickle Cell Trait. J. Hematol. 11, 92–108 (2022).

18. Pamba, A. et al. Clinical spectrum and severity of hemolytic anemia in glucose 6- phosphate dehydrogenase-deficient children receiving dapsone. Blood 120, 4123–4133 (2012).

19. Israel, A. et al. Health disparities in diabetes treatment: The challenge of G6PD deficiency. Diabetes Res. Clin. Pract. 219, 111965 (2025).

20. Madhani, A. et al. Clinical Penetrance of the Transthyretin V122I Variant in Older Black Patients With Heart Failure: The SCAN-MP (Screening for Cardiac Amyloidosis With Nuclear Imaging in Minority Populations) Study. J. Am. Heart Assoc. 12, e028973 (2023).

21. Jaiswal, S. et al. Clonal Hematopoiesis and Risk of Atherosclerotic Cardiovascular Disease. N. Engl. J. Med. 377, 111–121 (2017).

22. Bick, A. G. et al. Inherited causes of clonal haematopoiesis in 97,691 whole genomes. Nature 586, 763–768 (2020).

23. Cerezo, M. et al. The NHGRI-EBI GWAS Catalog: standards for reusability, sustainability and diversity. Nucleic Acids Res. 53, D998–D1005 (2025).

24. Sudlow, C. et al. UK biobank: an open access resource for identifying the causes of a wide range of complex diseases of middle and old age. PLoS Med. 12, e1001779 (2015).

25. Kurki, M. I. et al. FinnGen provides genetic insights from a well-phenotyped isolated population. Nature 613, 508–518 (2023).

26. Bastarache, L. et al. The phenotype-genotype reference map: Improving biobank data science through replication. Am. J. Hum. Genet. 110, 1522–1533 (2023).

27. Research, C. for D. E. and. FDA Adverse Event Reporting System (FAERS) Public Dashboard. FDA (2023).

28. Insani, W. N. et al. Prevalence of adverse drug reactions in the primary care setting: A systematic review and meta-analysis. PLoS ONE 16, e0252161 (2021).

29. Lazarou, J., Pomeranz, B. H. & Corey, P. N. Incidence of adverse drug reactions in hospitalized patients: a meta-analysis of prospective studies. JAMA 279, 1200–1205 (1998).

30. Brown, N. J., Ray, W. A., Snowden, M. & Griffin, M. R. Black Americans have an increased rate of angiotensin converting enzyme inhibitor-associated angioedema. Clin. Pharmacol. Ther. 60, 8–13 (1996).

31. Pirmohamed, M. et al. Adverse drug reactions as cause of admission to hospital: prospective analysis of 18 820 patients. BMJ 329, 15–19 (2004).

32. McCormack, M. et al. HLA-A*3101 and carbamazepine-induced hypersensitivity reactions in Europeans. N. Engl. J. Med. 364, 1134–1143 (2011).

33. Chung, W.-H. et al. Medical genetics: a marker for Stevens-Johnson syndrome. Nature 428, 486 (2004).

34. Sun, B. et al. Development and Application of Pharmacological Statin-Associated Muscle Symptoms Phenotyping Algorithms Using Structured and Unstructured Electronic Health Records Data. medRxiv 2023.05.04.23289523 (2023) doi:10.1101/2023.05.04.23289523.

35. Zheng, N. S. et al. High-throughput framework for genetic analyses of adverse drug reactions using electronic health records. PLoS Genet. 17, e1009593 (2021).

36. Soremekun, O. et al. Genome-Wide Association and Mendelian Randomization Analysis Reveal the Causal Relationship Between White Blood Cell Subtypes and Asthma in Africans. Front. Genet. 12, 749415 (2021).

37. Sun, Q. et al. Analyses of biomarker traits in diverse UK biobank participants identify associations missed by European-centric analysis strategies. J. Hum. Genet. 67, 87–93 (2022).

38. Verma, A. et al. Diversity and scale: Genetic architecture of 2068 traits in the VA Million Veteran Program. Science 385, eadj1182 (2024).

39. Bijlsma-Rutte, A., Rutters, F., Elders, P. J. M., Bot, S. D. M. & Nijpels, G. Socio- economic status and HbA1c in type 2 diabetes: A systematic review and meta-analysis. Diabetes Metab. Res. Rev. 34, e3008 (2018).

40. Hill-Briggs, F. et al. Social Determinants of Health and Diabetes: A Scientific Review. Diabetes Care 44, 258–279 (2020).

41. Walsh, C. G. et al. Scalable incident detection via natural language processing and probabilistic language models. Sci. Rep. 14, 23429 (2024).

42. Bejan, C. A. et al. Mining 100 million notes to find homelessness and adverse childhood experiences: 2 case studies of rare and severe social determinants of health in electronic health records. J. Am. Med. Inform. Assoc. JAMIA 25, 61–71 (2018).

43. Rebbeck, T. R., Mahal, B., Maxwell, K. N., Garraway, I. P. & Yamoah, K. The distinct impacts of race and genetic ancestry on health. Nat. Med. 28, 890–893 (2022).

44. Bryc, K., Durand, E. Y., Macpherson, J. M., Reich, D. & Mountain, J. L. The Genetic Ancestry of African Americans, Latinos, and European Americans across the United States. Am. J. Hum. Genet. 96, 37–53 (2015).

45. Benson, J. M. & Therrell, B. L. History and current status of newborn screening for hemoglobinopathies. Semin. Perinatol. 34, 134–144 (2010).

46. Ala, C. et al. A critical review of therapeutic interventions in sickle cell disease: Progress and challenges. Arch. Pharm. (Weinheim) 357, e2400381 (2024).

47. Kurian, A. W. et al. Germline Genetic Testing After Cancer Diagnosis. JAMA 330, 43– 51 (2023).

48. Guo, F., Scholl, M., Fuchs, E. L., Berenson, A. B. & Kuo, Y.-F. BRCA testing and testing results among women 18-65 years old. Prev. Med. Rep. 26, 101738 (2022).

49. Young, J., Bhattacharya, K., Ramachandran, S., Lee, A. & Bentley, J. P. Rates of genetic testing in patients prescribed drugs with pharmacogenomic information in FDA-approved labeling. Pharmacogenomics J. 21, 318–325 (2021).

50. Kurian, A. W. et al. Genetic Testing and Results in a Population-Based Cohort of Breast Cancer Patients and Ovarian Cancer Patients. J. Clin. Oncol. Off. J. Am. Soc. Clin. Oncol. 37, 1305–1315 (2019).

51. Chapman-Davis, E. et al. Racial and Ethnic Disparities in Genetic Testing at a Hereditary Breast and Ovarian Cancer Center. J. Gen. Intern. Med. 36, 35–42 (2021).

52. Regan, J. A. et al. Should We Systematically Screen for the Amyloidogenic V142I Variant? J. Card. Fail. 31, 136–139 (2025).

53. Hulsizer, J. et al. Sickle Cell Trait and Risk for Common Diseases: Evidence from the UK Biobank. Am. J. Med. 135, e279–e287 (2022).

54. Pershad, Y. et al. A clinical algorithm to identify people with the glucose-6-phosphate dehydrogenase p.Val68Met variant at risk for diabetes undertreatment. MedRxiv Prepr. Serv. Health Sci. 2025.01.17.25320736 (2025) doi:10.1101/2025.01.17.25320736.

55. Gibbs, C. R., Lip, G. Y. H. & Beevers, D. G. Angioedema due to ACE inhibitors: increased risk in patients of African origin. Br. J. Clin. Pharmacol. 48, 861–865 (1999).

56. Banerji, A., Blumenthal, K. G., Lai, K. H. & Zhou, L. Epidemiology and Incidence of ACE Inhibitor Angioedema Utilizing a Large Electronic Health Record. J. Allergy Clin. Immunol. Pract. 5, 744–749 (2017).

57. Brokamp, C. et al. Material community deprivation and hospital utilization during the first year of life: an urban population-based cohort study. Ann. Epidemiol. 30, 37–43 (2019).

58. Chakravarthy, R. et al. Determinants of stage at diagnosis of HPV-related cancer including area deprivation and clinical factors. J. Public Health Oxf. Engl. 44, 18–27 (2022).

59. Braveman, P. A., Arkin, E., Proctor, D., Kauh, T. & Holm, N. Systemic And Structural Racism: Definitions, Examples, Health Damages, And Approaches To Dismantling. Health Aff. Proj. Hope 41, 171–178 (2022).

60. Behera, S. et al. Comprehensive and accurate genome analysis at scale using DRAGEN accelerated algorithms. BioRxiv Prepr. Serv. Biol. 2024.01.02.573821 (2024) doi:10.1101/2024.01.02.573821.

61. Hofmeister, R. J., Ribeiro, D. M., Rubinacci, S. & Delaneau, O. Accurate rare variant phasing of whole-genome and whole-exome sequencing data in the UK Biobank. Nat. Genet. 55, 1243–1249 (2023).

62. Vlasschaert, C. et al. A practical approach to curate clonal hematopoiesis of indeterminate potential in human genetic data sets. Blood 141, 2214–2223 (2023).

63. Jaiswal, S. et al. Clonal Hematopoiesis and Risk of Atherosclerotic Cardiovascular Disease. N. Engl. J. Med. 377, 111–121 (2017).

64. Introducing Genomics in the Cloud, ie ‘The GATK book’. GATK https://gatk.broadinstitute.org/hc/en-us/articles/360042408831-Introducing-Genomics-in-the-Cloud-ie-The-GATK-book (2020).

65. Wang, K., Li, M. & Hakonarson, H. ANNOVAR: functional annotation of genetic variants from high-throughput sequencing data. Nucleic Acids Res. 38, e164 (2010).

66. 1000 Genomes Project Consortium et al. A global reference for human genetic variation. Nature 526, 68–74 (2015).

67. Zhou, W. et al. Global Biobank Meta-analysis Initiative: Powering genetic discovery across human disease. Cell Genomics 2, 100192 (2022).

68. Chang, C. C. et al. Second-generation PLINK: rising to the challenge of larger and richer datasets. GigaScience 4, 7 (2015).

69. Chiu, A. M., Molloy, E. K., Tan, Z., Talwalkar, A. & Sankararaman, S. Inferring population structure in biobank-scale genomic data. Am. J. Hum. Genet. 109, 727–737 (2022).

70. Anderson, C. A. et al. Data quality control in genetic case-control association studies. Nat. Protoc. 5, 1564–1573 (2010).

71. Koenig, Z. et al. A harmonized public resource of deeply sequenced diverse human genomes. Genome Res. 34, 796–809 (2024).

72. Maples, B. K., Gravel, S., Kenny, E. E. & Bustamante, C. D. RFMix: a discriminative modeling approach for rapid and robust local-ancestry inference. Am. J. Hum. Genet. 93, 278– 288 (2013).

73. Lisi, A. & Campbell, M. C. AncestryGrapher toolkit: Python command-line pipelines to visualize global- and local- ancestry inferences from the RFMIX version 2 software. Bioinforma. Oxf. Engl. 40, btae616 (2024).

74. Bastarache, L., Denny, J. C. & Roden, D. M. Phenome-Wide Association Studies. JAMA 327, 75–76 (2022).

75. Goldstein, B. A., Bhavsar, N. A., Phelan, M. & Pencina, M. J. Controlling for Informed Presence Bias Due to the Number of Health Encounters in an Electronic Health Record. Am. J. Epidemiol. 184, 847–855 (2016).

76. Dennis, J. K. et al. Clinical laboratory test-wide association scan of polygenic scores identifies biomarkers of complex disease. Genome Med. 13, 6 (2021).

77. Shuey, M. M., et al. Next-generation phenotyping: introducing phecodeX for enhanced discovery research in medical phenomics. Bioinforma. Oxf. Engl. 39, btad655 (2023).

78. Dorr, D. et al. Identifying Patients with Significant Problems Related to Social Determinants of Health with Natural Language Processing. Stud. Health Technol. Inform. 264, 1456–1457 (2019).

79. Bejan, C. A. et al. Improving ascertainment of suicidal ideation and suicide attempt with natural language processing. Sci. Rep. 12, 15146 (2022).

80. Jasra, S. et al. High burden of clonal hematopoiesis in first responders exposed to the World Trade Center disaster. Nat. Med. 28, 468–471 (2022).

81. Ge, T., Chen, C.-Y., Ni, Y., Feng, Y.-C. A. & Smoller, J. W. Polygenic prediction via Bayesian regression and continuous shrinkage priors. Nat. Commun. 10, 1776 (2019).

82. Zheng, N. S. et al. High-throughput framework for genetic analyses of adverse drug reactions using electronic health records. PLoS Genet. 17, e1009593 (2021).

