## Supplementary Figures for "Genome sequencing of 35,024 predominantly African ancestry persons addresses gaps in genomics and healthcare"

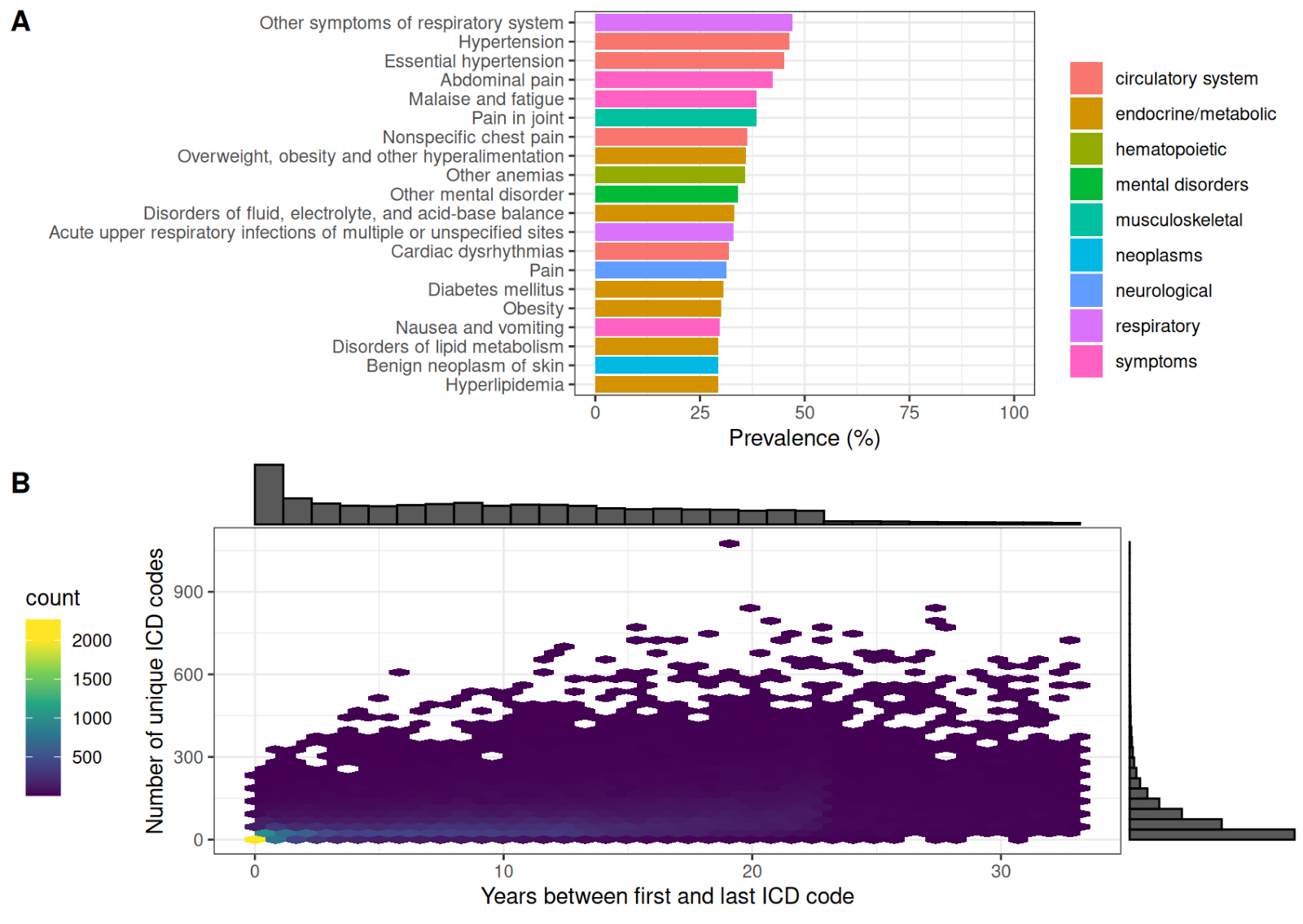

**Figure S1:** A) Top ten most prevalent phecodes in the AGD35k cohort and B) distribution of the length between the first and last ICD code and the number of unique ICD codes for each participant.

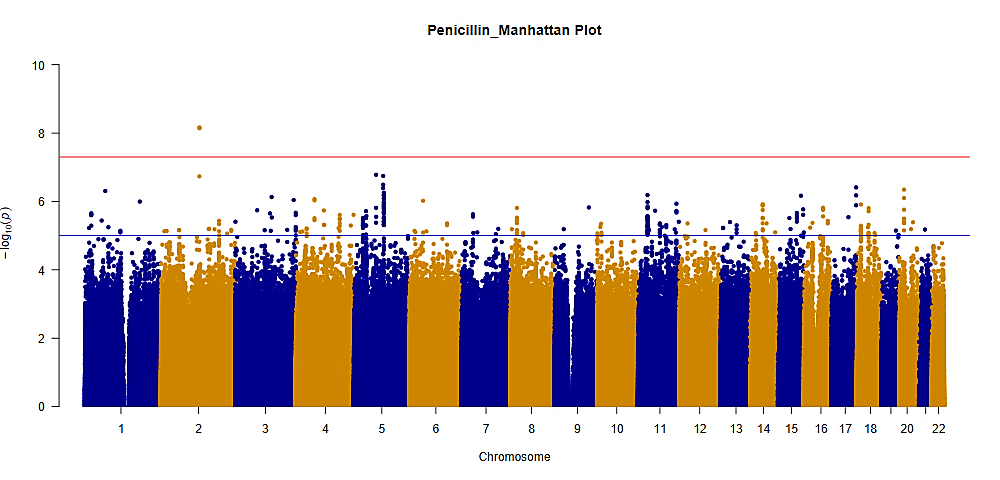
A)

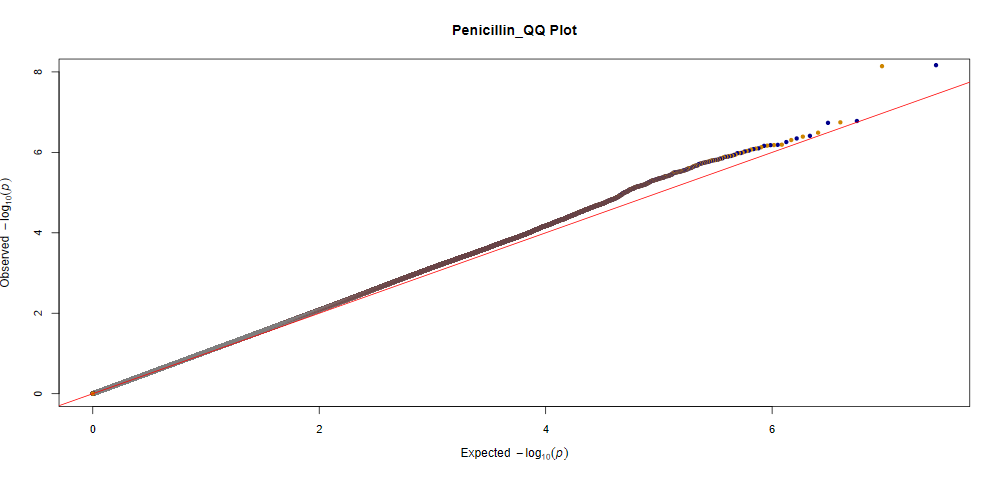
B)

GIF:1.05

**Figure S2.** A) Manhattan plot for the GWAS of penicillin adverse drug events. SNPs were plotted based on their physical chromosomal positions (horizontal axis) together with their-log 10 (P-values) in the GWAS (vertical axis). The red horizontal line shows the genome-wide significance threshold of P=5.0x10^-8^. The blue horizontal line shows the threshold of P=1.0x10^-5^. B) Quantile-quantile plot for penicillin adverse drug events. Expected P-values assuming a uniform distribution (X-axis; expected 2log10(P-value)) were compared to observed p-values (Y-axis; observed 2log10(P-value)). GIF = genomic inflation factor.

A)

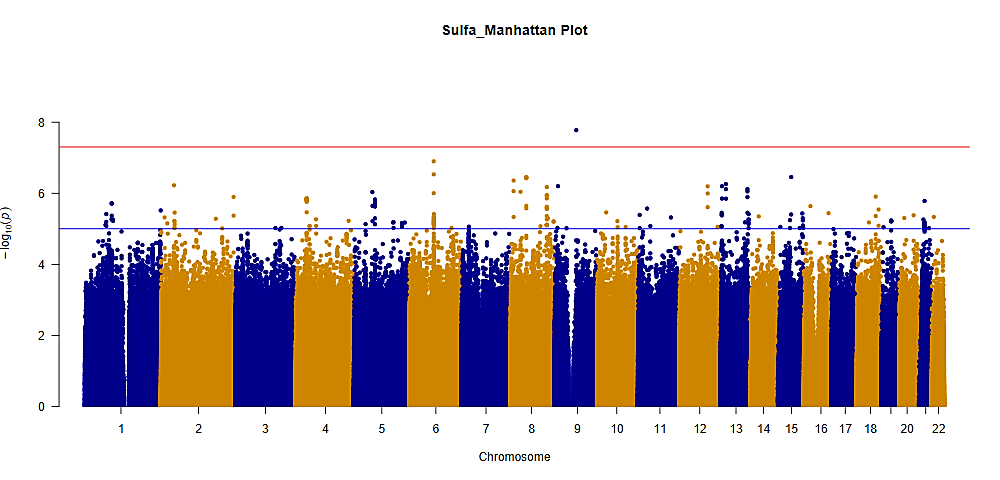

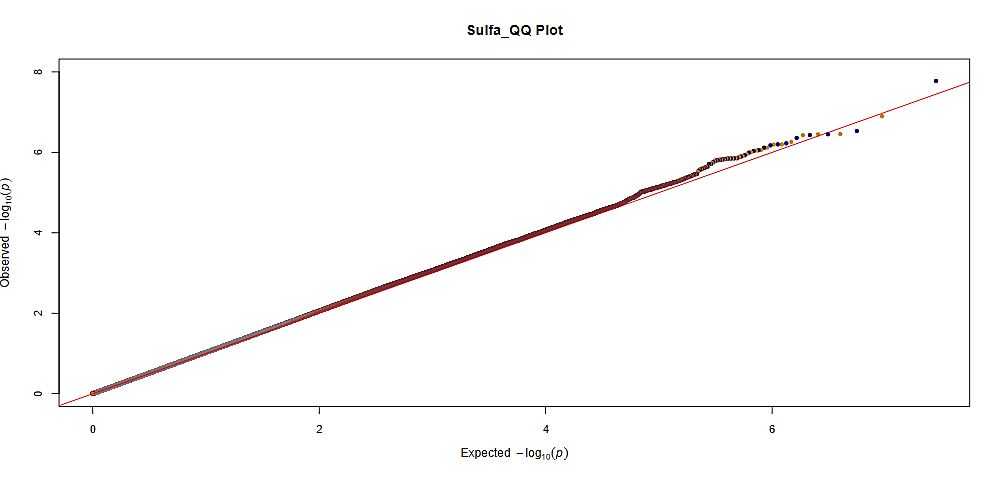
B)

GIF: 1.03

**Figure S3.** A) Manhattan plot for the GWAS of sulfa adverse drug events. SNPs were plotted based on their physical chromosomal positions (horizontal axis) together with their-log 10 (P-values) in the GWAS (vertical axis). The red horizontal line shows the genome-wide significance threshold of P=5.0x10-8. The blue horizontal line shows the threshold of P=1.0x10-5. B) Quantile-quantile plot for sulfa adverse drug events. Expected P-values assuming a uniform distribution X-axis; expected 2log10(P-value)) were compared to observed p-values (Y-axis; observed 2log10(P-value)). GIF = genomic inflation factor.

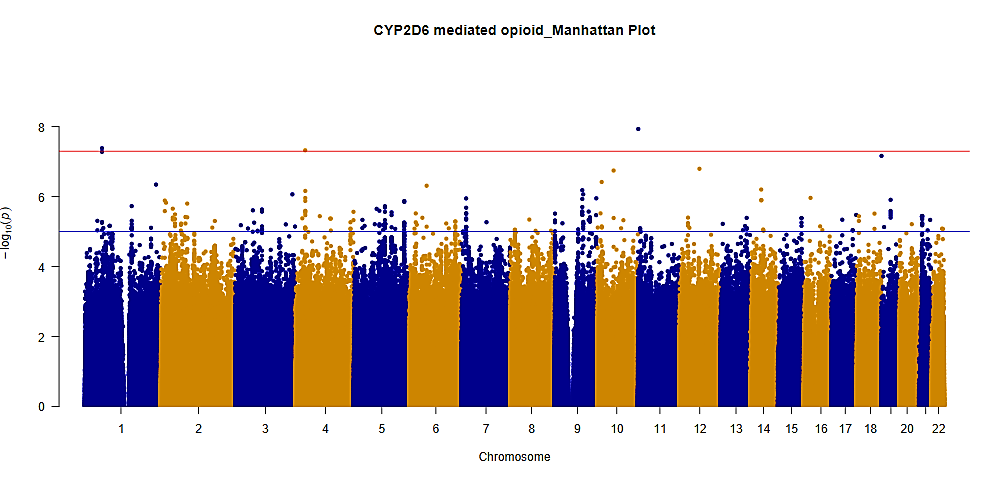
A)

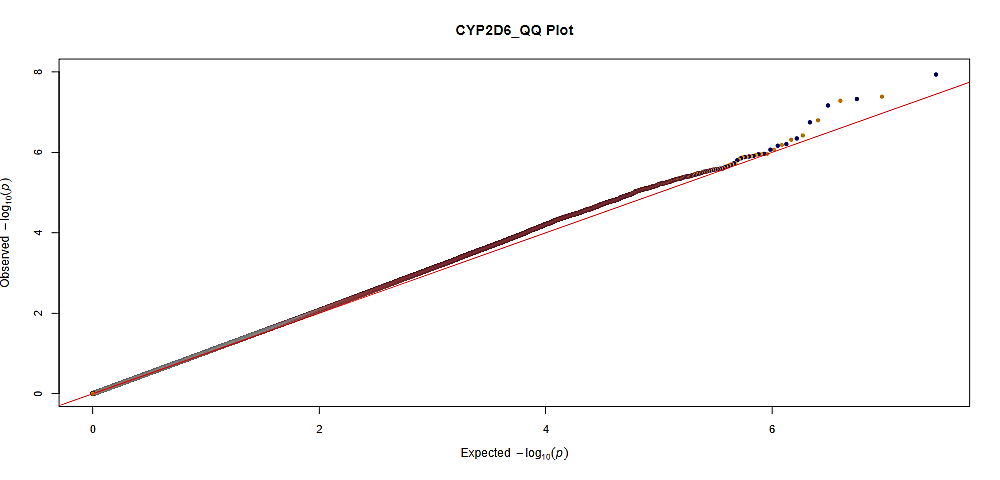
B)

GIF: 1.05

**Figure S4.** A) Manhattan plot for the GWAS of CYP2D6-mediated opioids adverse drug events. SNPs were plotted based on their physical chromosomal positions (horizontal axis) together with their-log 10 (P-values) in the GWAS (vertical axis). The red horizontal line shows the genome-wide significance threshold of P=5.0x10-8. The blue horizontal line shows the threshold of P=1.0x10-5. B) Quantile-quantile plot for CYP2D6-mediated opioids adverse drug events. Expected P-values assuming a uniform distribution X-axis; expected 2log10(P-value)) were compared to observed p-values (Y-axis; observed 2log10(P-value)). GIF = genomic inflation factor.

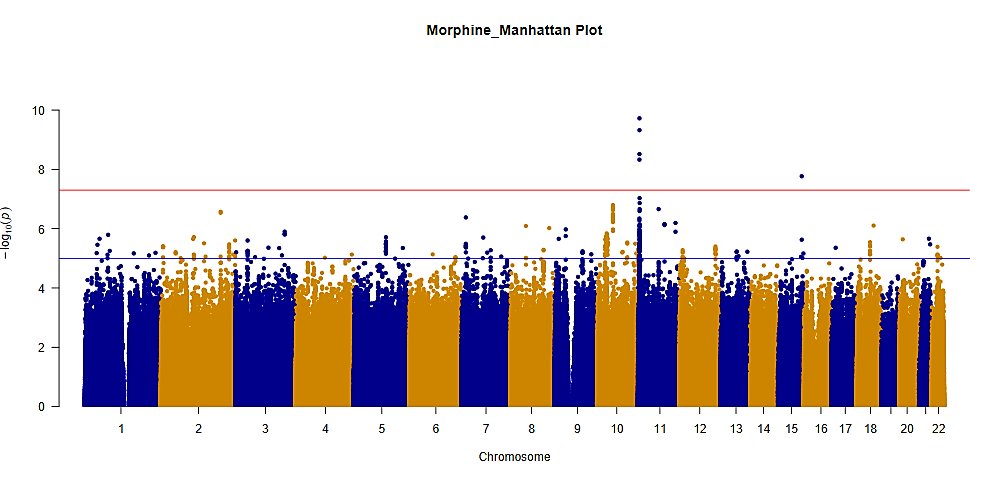
A)

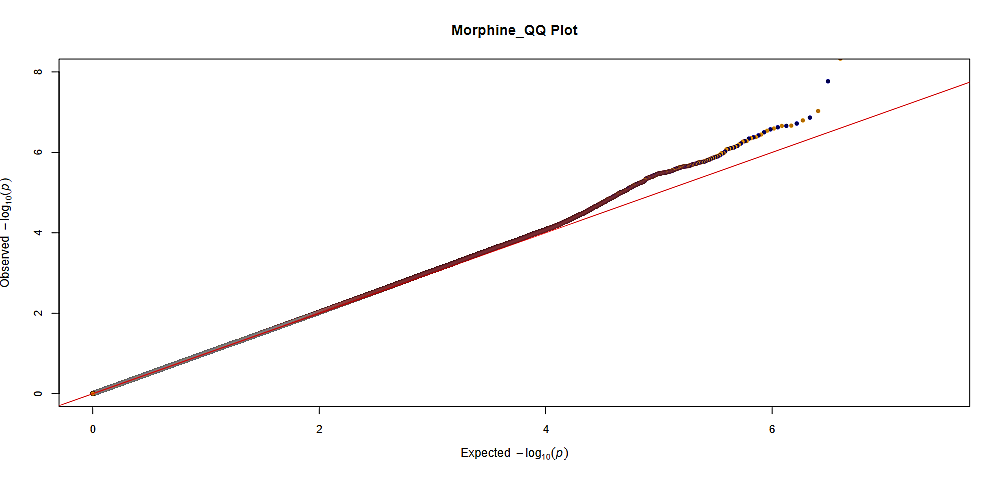
B)

GIF: 1.02

**Figure S5.** A) Manhattan plot for the GWAS of morphine adverse drug events. SNPs were plotted based on their physical chromosomal positions (horizontal axis) together with their-log 10 (P-values) in the GWAS (vertical axis). The red horizontal line shows the genome-wide significance threshold of P=5.0x10-8. The blue horizontal line shows the threshold of P=1.0x10-5. B) Quantile-quantile plot for morphine adverse drug events. Expected P-values assuming a uniform distribution X-axis; expected 2log10(P-value)) were compared to observed p-values (Y-axis; observed 2log10(P-value)). GIF = genomic inflation factor.

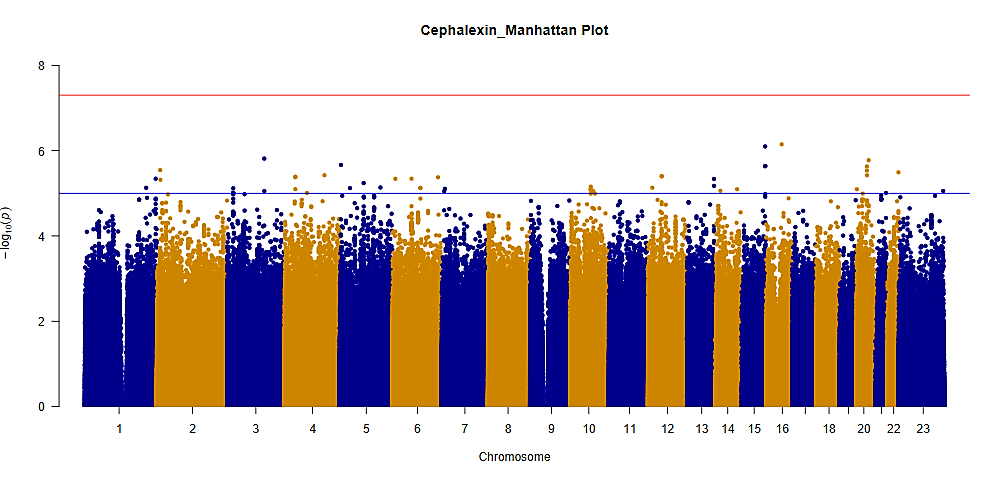
A)

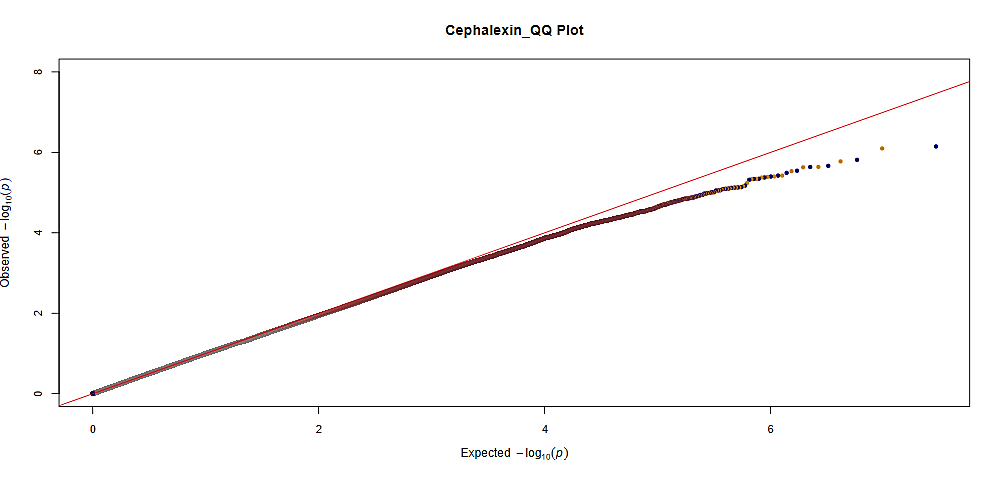
B)

GIF: 1.01

**Figure S6.** A) Manhattan plot for the GWAS of cephalexin adverse drug events. SNPs were plotted based on their physical chromosomal positions (horizontal axis) together with their-log 10 (P-values) in the GWAS (vertical axis). The red horizontal line shows the genome-wide significance threshold of P=5.0x10-8. The blue horizontal line shows the threshold of P=1.0x10-5. B) Quantile-quantile plot for cephalexin adverse drug events. Expected P-values assuming a uniform distribution X-axis; expected 2log10(P-value)) were compared to observed p-values (Y-axis; observed 2log10(P-value)). GIF = genomic inflation factor.

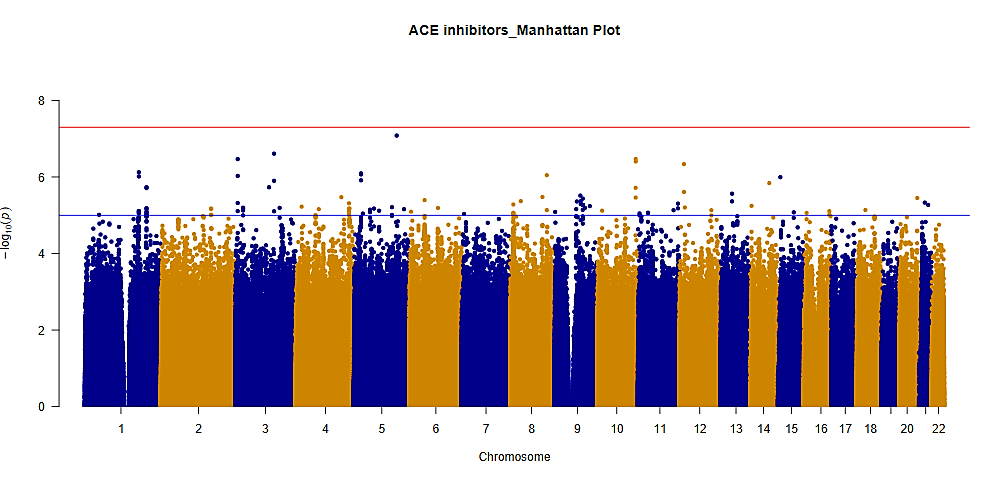
A)

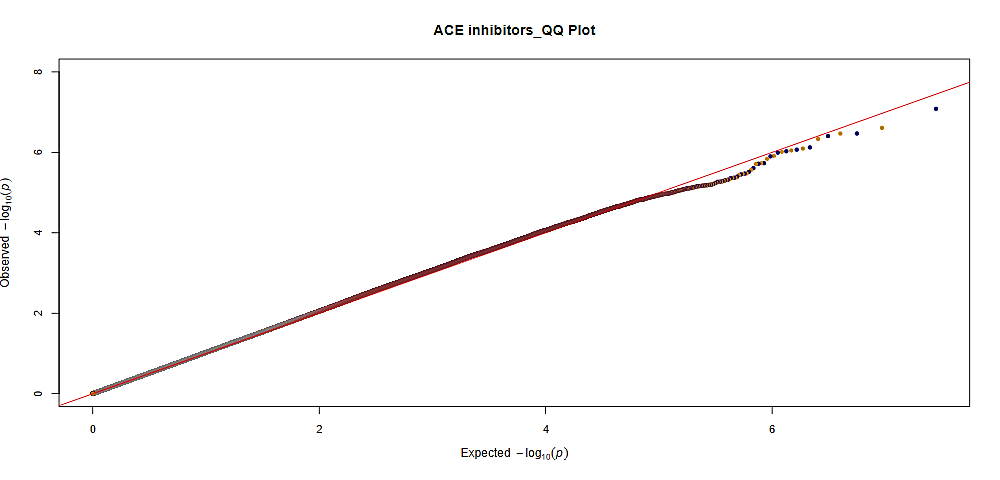
B)

GIF: 1.03

**Figure S7.** A) Manhattan plot for the GWAS of angiotensin-converting enzyme (ACE) inhibitor adverse drug events. SNPs were plotted based on their physical chromosomal positions (horizontal axis) together with their-log 10 (P-values) in the GWAS (vertical axis). The red horizontal line shows the genome-wide significance threshold of P=5.0x10-8. The blue horizontal line shows the threshold of P=1.0x10-5. B) Quantile-quantile plot for ACE inhibitor adverse drug events. Expected P-values assuming a uniform distribution X-axis; expected 2log10(P-value)) were compared to observed p-values (Y-axis; observed 2log10(P-value)). GIF = genomic inflation factor.

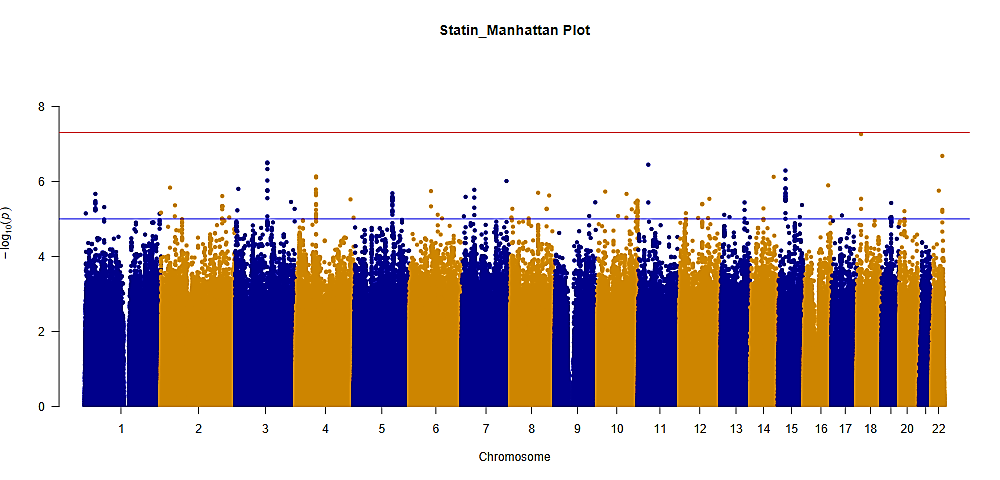
A)

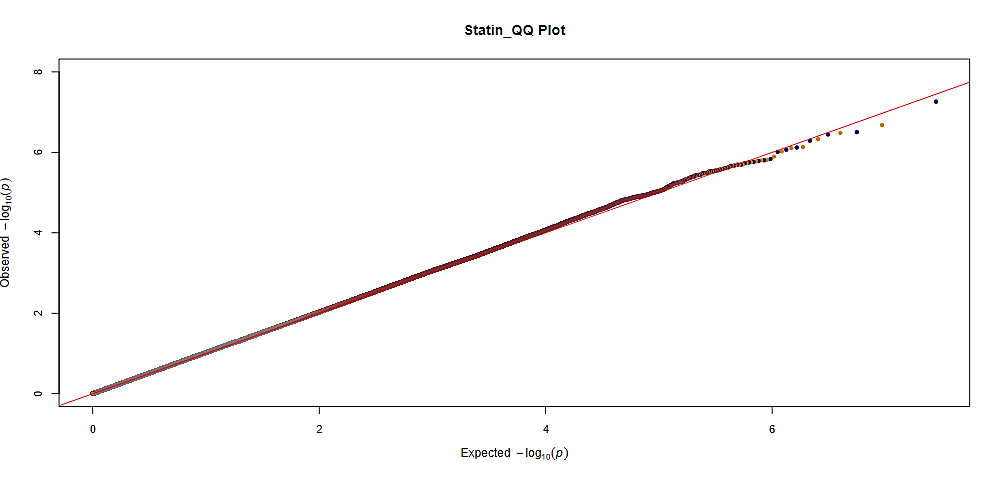
B)

GIF: 1.04

**Figure S8.** A) Manhattan plot for the GWAS of statin adverse drug events. SNPs were plotted based on their physical chromosomal positions (horizontal axis) together with their-log 10 (P-values) in the GWAS (vertical axis). The red horizontal line shows the genome-wide significance threshold of P=5.0x10-8. The blue horizontal line shows the threshold of P=1.0x10-5. B) Quantile-quantile plot for statin adverse drug events. Expected P-values assuming a uniform distribution X-axis; expected 2log10(P-value)) were compared to observed p-values (Y-axis; observed 2log10(P-value)). GIF = genomic inflation factor.

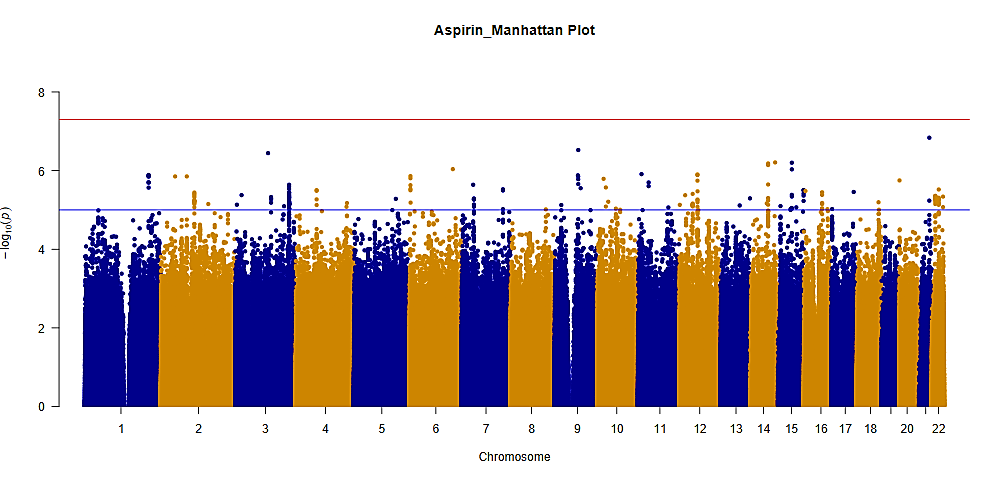
A)

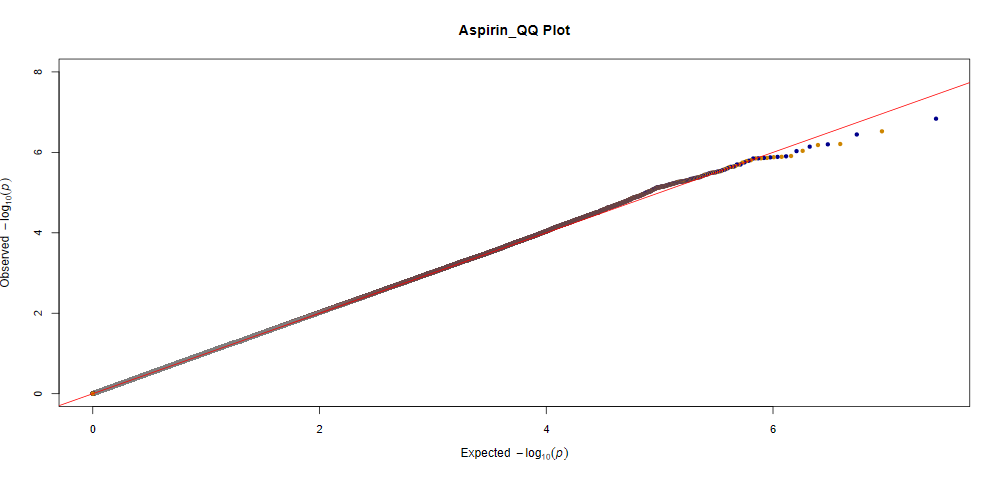
B)

GIF: 1.03

**Figure S9.** A) Manhattan plot for the GWAS of aspirin adverse drug events. SNPs were plotted based on their physical chromosomal positions (horizontal axis) together with their-log 10 (P-values) in the GWAS (vertical axis). The red horizontal line shows the genome-wide significance threshold of P=5.0x10-8. The blue horizontal line shows the threshold of P=1.0x10-5. B) Quantile-quantile plot for aspirin adverse drug events. Expected P-values assuming a uniform distribution X-axis; expected 2log10(P-value)) were compared to observed p-values (Y-axis; observed 2log10(P-value)). GIF = genomic inflation factor.

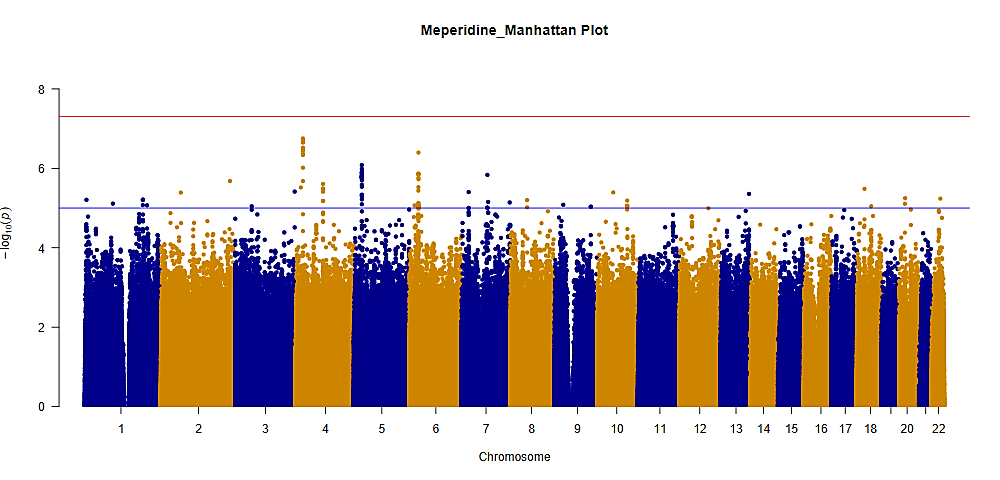
A)

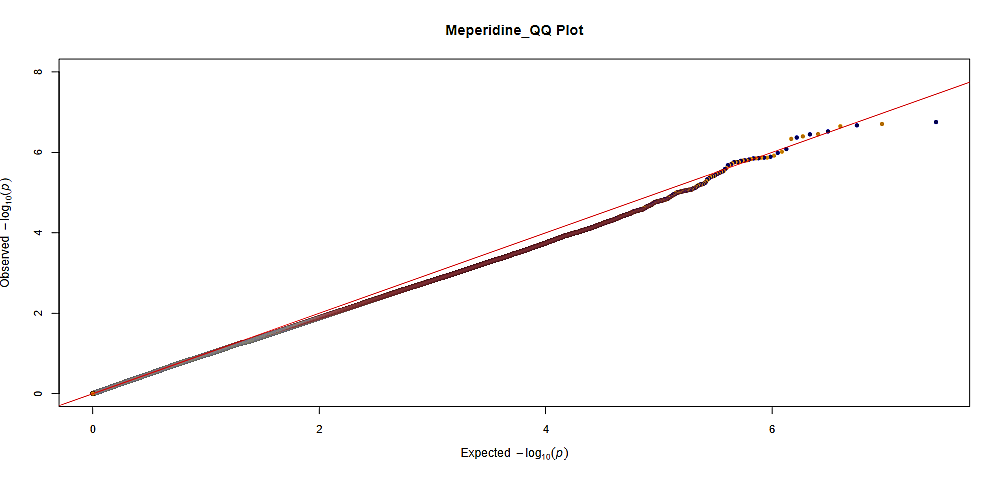
B)

GIF: 0.99

**Figure S10.** A) Manhattan plot for the GWAS of meperidine adverse drug events. SNPs were plotted based on their physical chromosomal positions (horizontal axis) together with their-log 10 (P-values) in the GWAS (vertical axis). The red horizontal line shows the genome-wide significance threshold of P=5.0x10-8. The blue horizontal line shows the threshold of P=1.0x10-5. B) Quantile-quantile plot for meperidine adverse drug events. Expected P-values assuming a uniform distribution X-axis; expected 2log10(P-value)) were compared to observed p-values (Y-axis; observed 2log10(P-value)). GIF = genomic inflation factor.

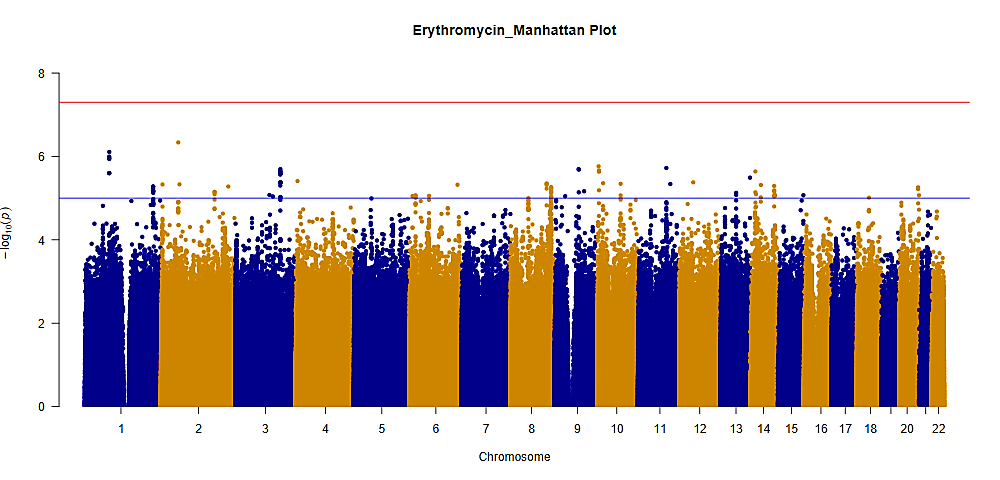
A)

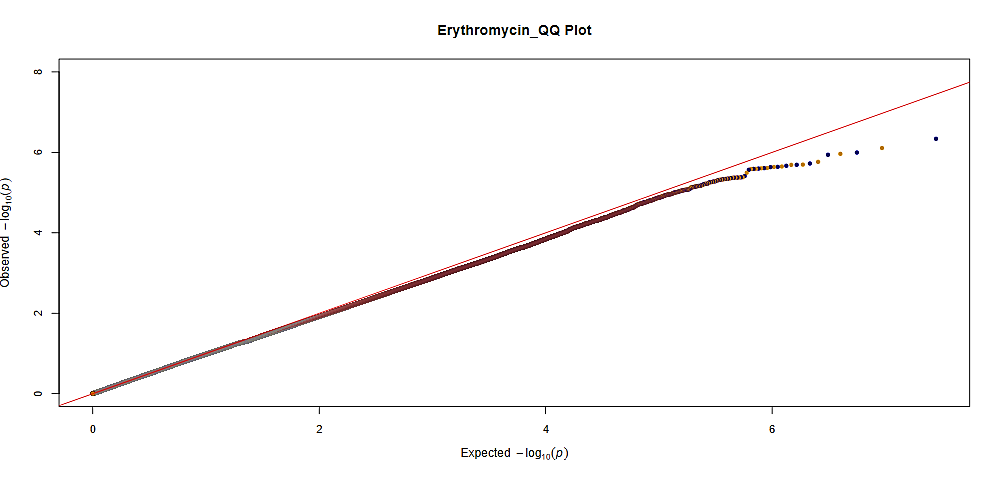
B)

GIF: 1.00

**Figure S11.** A) Manhattan plot for the GWAS of erythromycin adverse drug events. SNPs were plotted based on their physical chromosomal positions (horizontal axis) together with their-log 10 (P-values) in the GWAS (vertical axis). The red horizontal line shows the genome-wide significance threshold of P=5.0x10-8. The blue horizontal line shows the threshold of P=1.0x10-5. B) Quantile-quantile plot for erythromycin adverse drug events. Expected P-values assuming a uniform distribution X-axis; expected 2log10(P-value)) were compared to observed p-values (Y-axis; observed 2log10(P-value)). GIF = genomic inflation factor.

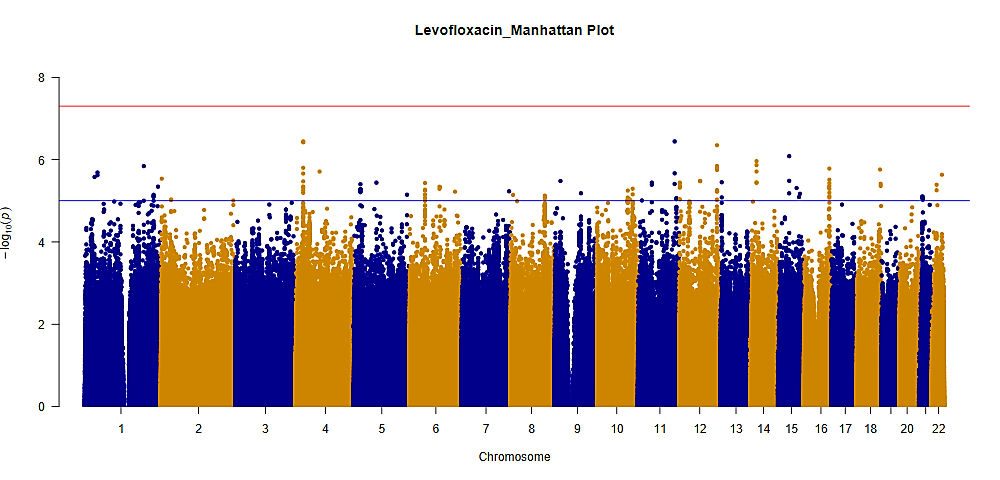
A)

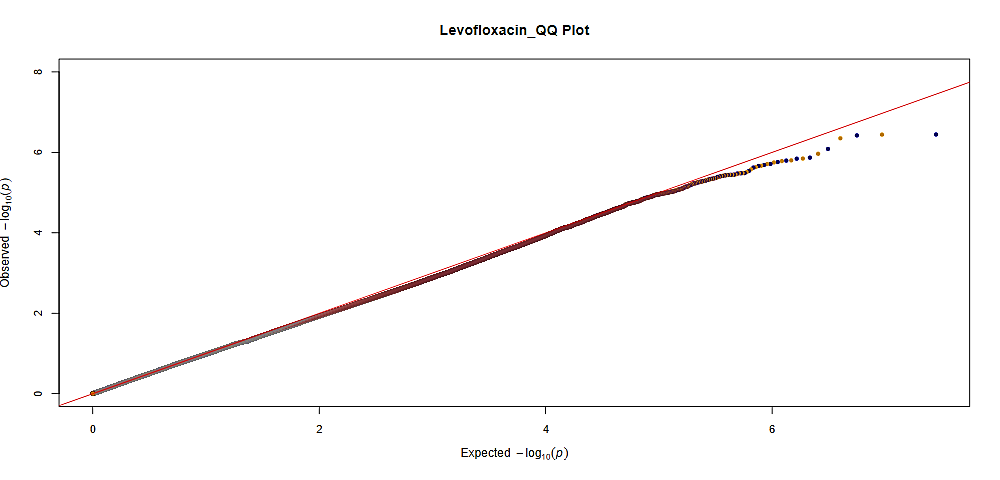
B)

GIF: 1.01

**Figure S12.** A) Manhattan plot for the GWAS of levofloxacin adverse drug events. SNPs were plotted based on their physical chromosomal positions (horizontal axis) together with their-log 10 (P-values) in the GWAS (vertical axis). The red horizontal line shows the genome-wide significance threshold of P=5.0x10-8. The blue horizontal line shows the threshold of P=1.0x10-5. B) Quantile-quantile plot for levofloxacin adverse drug events. Expected P-values assuming a uniform distribution X-axis; expected 2log10(P-value)) were compared to observed p-values (Y-axis; observed 2log10(P-value)). GIF = genomic inflation factor.
