## Supplementary Information for "Genome sequencing of 35,024 predominantly African ancestry persons addresses gaps in genomics and healthcare"

**Supplementary Results**

**Characterization of relatedness**

To better understand the generational depth of inherited ancestral segments, we estimated relatedness, predicted likely pedigrees, and identified shared genomic segments in 34,523 participants from the AGD35k cohort (see Methods). This analysis was completed prior to complete sequencing of the AGD35k cohort; therefore, 501 participants are missing from this analysis. We determined pair-wise close relatedness (1^st^ and 2^nd^ degree) and defined pedigrees using COMPADRE and we also determined pair-wise distant relatedness (3^rd^ degree and higher) using ERSA^1,2^. We identified ~627 million total identity by descent (IBD) segments shared pairwise with length equal to or greater than 2cM among 247,967,491 million pairs of participants. We detected 4,303 1^st^ degree relative pairs (parent-offspring or siblings), 4,673 2^nd^ degree relative pairs, and 13,083 3^rd^ degree relative pairs. There were 74,638 pairs related at 4^th^ or 5^th^ degree, 299,435 pairs related at 6^th^ or 7^th^, and over six million pairs related at 8^th^ or 9^th^ degree. We reconstructed 4,620 pedigrees for participants related at 3^rd^ degree and closer. Of these pedigrees, 62% had two members, 19% had three members, 13% had four to five members, 5% had six to ten members, and <1% of pedigrees had greater than 10 members. The largest pedigree contained 69 individuals.

**Phenome-wide association study of chr17q21.3 inversion**

From the class of structural variants, we assessed the prevalence and consequence of a well-documented 900kb inversion at 17q21.3, observed almost exclusively in European and Middle Eastern genetic ancestries^3^. The inversion has previously been associated with a wide range of conditions including Parkinson’s disease, Progressive Supranuclear Palsy (PSP), blood cell composition, and reduced lung function^4^. Using 10 previously validated marker SNPs^5^, we tagged participants in the AGD35k cohort as possessing 0, 1, or 2 copies of the inversion. The inversion was present in 13% AGD35k participants, compared to 38% in the UK biobank (UKB). Given the hypothesized European origins of this inversion^3,6^, we assessed the relationship between European admixture and inversion carrier frequency. For each continental ancestry group, we quantified the mean proportion of European genetic ancestry in inversion carriers against non-carriers. We observed statistically significant differences in four of the five populations. The direction of the relationships consistently favored higher proportion of European genetic ancestry among inversion carriers.

Phenome wide association studies (PheWAS) of the inversion did not identify statistically significant results in either European or African ancestry cohorts. Previous PheWAS of the inversion have been conducted by screening the GWAS catalog for associations with a single inversion tagging SNP and reporting the most significant phenotypes^4,7,8^. An analysis using SNP profiling across the inversion region in the UKB identified associations with hip arthrosis, hemorrhoids, Parkinson’s disease, digit deformities, anal fissures, and pulmonary diseases^9^. We were unable to replicate the accompanying phecode associations in the subset of AGD35k with >50% African genetic ancestry.

**Supplementary Methods**

**Inversion tagging of chr17q21.3**

The presence of the chr17q21.3 inversion was assessed using SNP-based tagging of the inversion-carrying H2 haplotype. We identified 12 inversion-tagging SNPs from the literature that demonstrate perfect linkage disequilibrium with the H2 haplotype in independent populations. Because these reference populations are of European descent, it is possible that the tag SNPs may have arisen independently outside the context of the inversion in non-European populations. We assessed the frequency of these tag SNPs in populations of African genetic ancestries using the 1000 genomes reference data set. Prior work has estimated the frequency of the inversion in these populations as below 2%. Accordingly, we excluded tag SNPs with allele frequencies greater than this threshold.

We used whole genome sequencing data to identify the number of inversion-tagging SNPs carried by each participant in the AGD35k population. We classify inversion carriers as those participants with an inversion-tagging SNP at ≥ 80% of the tested loci.

**PheWAS of the chr17q21.3 inversion**

We performed a PheWAS of the chr17q21.3 inversion in parallel between cohorts from the AGD35k and the UKB. These PheWAS included phecodes, aggregates of clinical billing codes, as the outcome variables and the chr17q21.3 inversion as the predictor. We used our muti-SNP marker method to classify participants according to the number of H2 inversion haplotypes present (0, 1, or 2). Analyses were adjusted for standard covariates including age, age-squared, and the first 20 principal components (PCs). We subset the AGD cohort into subcategories of individuals with >50% African ancestry and individuals with >50% European ancestry for each analysis. Bonferroni correction was used to adjust for multiple testing.

**Supplementary References**

1.	Staples, J. *et al.* PADRE: Pedigree-Aware Distant-Relationship Estimation. *Am. J. Hum. Genet.* **99**, 154–162 (2016).

2.	Huff, C. D. *et al.* Maximum-likelihood estimation of recent shared ancestry (ERSA). *Genome Res.* **21**, 768–774 (2011).

3.	Stefansson, H. *et al.* A common inversion under selection in Europeans. *Nat. Genet.* **37**, 129–137 (2005).

4.	Campoy, E., Puig, M., Yakymenko, I., Lerga-Jaso, J. & Cáceres, M. Genomic architecture and functional effects of potential human inversion supergenes. *Philos. Trans. R. Soc. B* (2022) doi:10.1098/rstb.2021.0209.

5.	Steinberg, K. M. *et al.* Structural diversity and African origin of the 17q21.31 inversion polymorphism. *Nat. Genet.* **44**, 872–880 (2012).

6.	Porubsky, D. *et al.* Recurrent inversion polymorphisms in humans associate with genetic instability and genomic disorders. *Cell* **185**, 1986-2005.e26 (2022).

7.	Pedicone, C., Weitzman, S. A., Renton, A. E. & Goate, A. M. Unraveling the complex role of MAPT-containing H1 and H2 haplotypes in neurodegenerative diseases. *Mol. Neurodegener.* **19**, 43 (2024).

8.	Puig, M. *et al.* Determining the impact of uncharacterized inversions in the human genome by droplet digital PCR. *Genome Res.* **30**, 724–735 (2020).

9.	Sergouniotis, P. I. *et al.* Autoencoder-based phenotyping of ophthalmic images highlights genetic loci influencing retinal morphology and provides informative biomarkers. *Bioinformatics* **41**, btae732 (2025).
